# COVID-19 pandemic surges can induce bias in trials using response adaptive randomization: A simulation study

**DOI:** 10.1101/2022.07.13.22277596

**Authors:** Christopher J Yarnell, Robert A Fowler, Lillian Sung, George Tomlinson

## Abstract

Response-adaptive randomization is being used in COVID-19 trials, but it is unknown whether outcome rate changes during surges of COVID-19 will lead to bias in trial results. In response-adaptive randomization, allocation ratios are adjusted according to interim analyses to assign more patients to promising interventions. Although it is known that response-adaptive randomization may give biased estimates if outcome rates drift over time, observed mortality fluctuations in the COVID-19 pandemic are more extreme than any previously tested in simulation. We hypothesized that pandemic surges induce bias in trials using response-adaptive randomization, and that adjustment for time will alleviate that bias. Bayesian 4-arm superiority trials with a mortality outcome were simulated to investigate bias in treatment effect, comparing complete and response-adaptive randomization under different pandemic scenarios based on data from New York, Spain, and Italy. Relative bias in the odds ratio associated with treatment ranged from 0.3% to 11% and was largest in trials with a surge and an effective intervention that did not adjust for time. Bias was attenuated by adjustment for time without compromising the false-positive rate. Trials using response-adaptive randomization were more likely to identify effective interventions but were slower to drop ineffective interventions. Even with variation in outcome rates similar to observed pandemic surges, COVID-19 trials using response-adaptive randomization that adjust for time can provide accurate estimates of treatment effects.

## Introduction

The SARS-CoV-2 pandemic triggered a global effort to test interventions in clinical trials.^1^ Some of the proposed and ongoing trials employ response-adaptive randomization, defined as a method of changing allocation to interventions in a randomized trial according to trial outcomes.^1–4^ This is a method of allocating more patients to effective interventions and potentially reducing the sample size required to reach a conclusion ^5–8^, but uptake has so far been uncommon.^9–11^

Response-adaptive randomization (RAR), as compared to complete (conventional) randomization, is potentially suitable for COVID-19 trials because RAR generally increases the probability that patients are allocated to effective arms. This may appeal to patients considering enrollment, clinicians screening patients or joining trials, and regulators assigning trial funding.^5,6,12–19^ In the setting of 2 or more interventions in addition to control there may also be a decrease in sample size required to identify successful therapies, depending on the method of adaptive randomization.^14,20–23^. Trials that use RAR may face logistical challenges ^19^, require complex analyses limiting the accessibility of results ^24^, and give biased estimates if the risk of outcomes changes over time. ^8,16,21,22,25^ This last limitation is particularly relevant to COVID-19. For example, if COVID-19 cases have overburdened the healthcare system, the mortality rate may increase and then return to baseline as the surge abates. If the allocation ratios of patients to different treatment arms are different during the time of high as opposed to baseline mortality, a naïve comparison of outcomes by treatment assignment could create the illusion of harm or efficacy between equivalent treatments (Figure 1).

**Figure 1:**
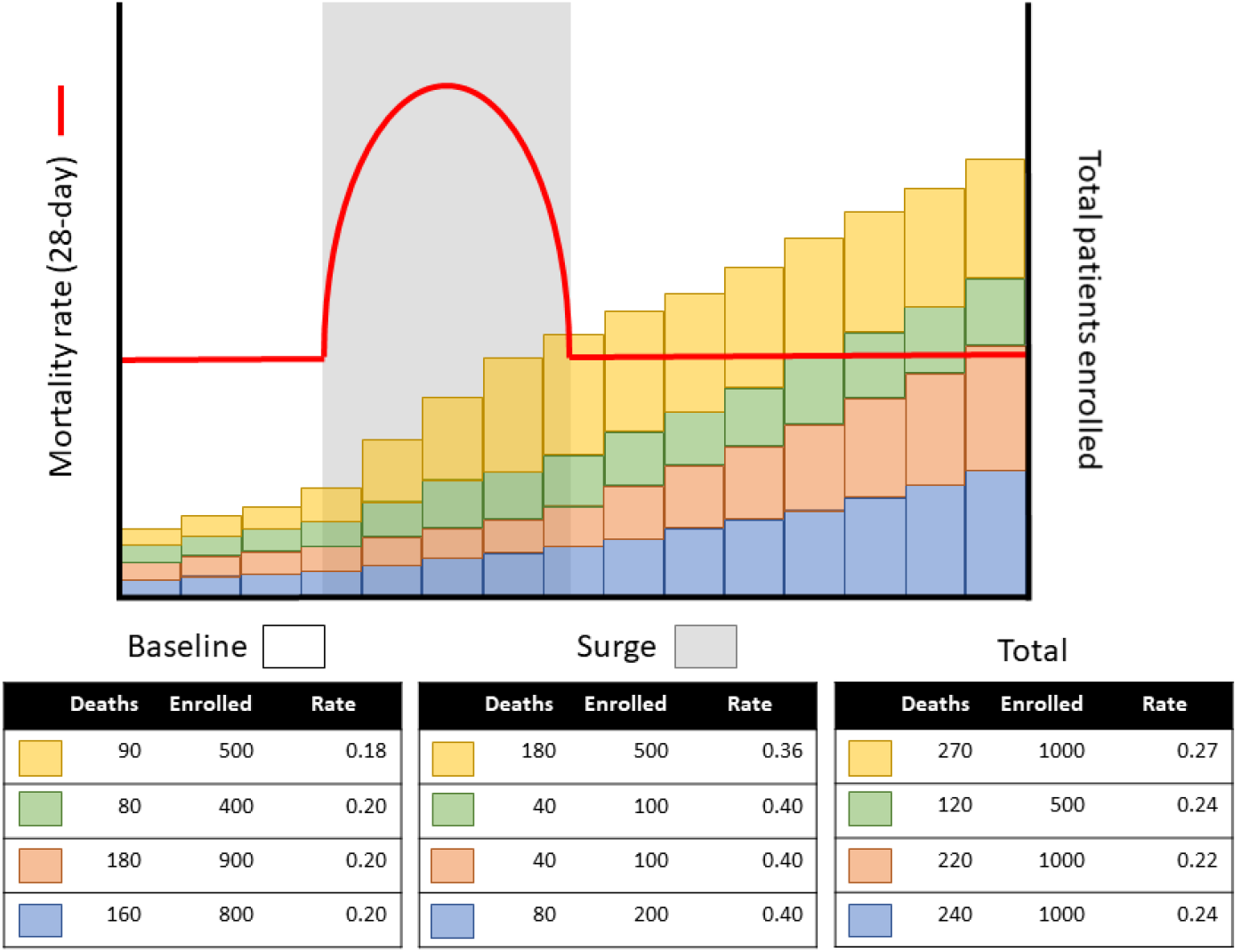
Potential bias due to adaptive randomization in a pandemic. This figure shows the 28-day mortality (red line) and number of patients allocated to each ofthe three treatment groups (yellow, green, red) and control (blue) versus time. The 28-day mortality rate doubles during a surge (translucent gray box). The tables below show the counts of patients enrolled either during baseline or surge conditions, with a third table showing the totals. The yellow intervention enrolls many more patients than the other treatments during the surge, so an unadjusted comparison suggests that the yellow group has a higher mortality rate (0.27) even though within either baseline or surge intervals it is beneficial (OR 0.88). By contrast, the red intervention appears to be beneficial because during the surge it was allocated the fewest patients, but within each interval it is no different from control (OR 1.0). Using logistic regression with time divided into intervals (visualized as blocks on the figure) provides a way of comparing patient outcomes within each time “block”.

COVID-19 outbreaks with surges in case numbers have occurred worldwide, causing shortages of medications and equipment, provision of intensive care in non-traditional settings, and wide variation in mortality rate for critically ill and hospitalized patients.^26–31^ The performance of adaptive randomization has been assessed in trials with linear drift, including drift based on the Ebola virus disease epidemiology.^12,32^ No work, to our knowledge, assesses the robustness of response-adaptive randomization to the nonlinear surges demonstrated by COVID-19.

### Aims

Use simulated trials of therapies for COVID-19 to: (1) compare bias in the estimated odds ratios between trials using response-adaptive and complete randomization; (2) assess whether using regression to adjust for time reduces the potential bias with RAR; and (3) identify whether RAR demonstrates other advantages in the setting of pandemic epidemiology.

## Methods

This simulation study followed recommendations of Morris et. al ^33^ and the STRESS simulation reporting guidelines ^34^. This research did not require research ethics board review according to the Tri-Council Policy Statement Article 2.1. ^35^ In this study we use a Bayesian statistical framework, similar to most contemporary trials that use RAR ^10,11,36^, although response-adaptive randomization can also be implemented using frequentist approaches ^6,32,37^. Additional details about the methods are available in the Supplement.

### Data-generating mechanism

Simulations generated randomized superiority trials of hospitalized patients with severe COVID-19 pneumonia comparing three interventions to control with a binary 28-day mortality outcome. Each trial ran for 365 days. Mortality was simulated as a binary random variable based on mortality rate, and daily enrollment was simulated as a Poisson random variable based on the mean daily enrollment. Important components of the data-generating mechanism detailed below were pandemic epidemiology, number and efficacy of intervention arms, randomization algorithms, statistical treatment of time-varying risk, and stopping rules.

### Pandemic epidemiology

Pandemic epidemiology impacted the mortality and enrollment rates. Based on available information we set the baseline 28-day mortality rate to 20%. ^26,38^ Surges compromising healthcare capacity have been observed in China ^39^, Italy ^27^, New York ^26^, Spain ^31^, and Brazil ^40^. Narrative reports describe provision of intensive care throughout the hospital during surge conditions, so we set the surge mortality rate to be similar to the intensive care unit mortality at 40%. ^41,42^ For enrollment rates we used information from completed and in-progress COVID-19 trials and set mean daily enrolment to be 15 patients at baseline and 25 patients during surge.^2,43,44^

The duration of each surge was set to 21 days, based on the large dataset from the United Kingdom.^42^ The first surge, when present, began on trial day 30. The cyclic surge scenario used three surges (spring, fall, winter) starting on trial day 30, 150, and 270, approximately similar to the 1918 pandemic.^45^ The base case had no surge, and alternative cases had one or three surges. Single surge cases with later surge timing (days 150 and 270) were investigated in sensitivity analyses.

### Number and efficacy of intervention arms

The trials evaluated three intervention arms and one control arm, mimicking large trials of COVID-19 evaluating multiple arms. ^2,43,46^ Preliminary results from COVID-19 trials suggested an odds ratio of 0.75 for benefit was feasible.^43,47^ We defined the “null” scenario as all interventions being equivalent (OR = 1) and the “nugget” scenario as one intervention effective (OR 0.75) and the remainder equivalent (OR = 1). The control arm was intervention 1 and if there was an effective intervention, it was intervention 2. As sensitivity analyses we conducted simulations where a single intervention was harmful with an odds ratio of 1.33 and where there was one effective, one harmful, and one equivalent intervention.^44^

### Randomization algorithms

The base case for randomization was complete randomization where patients have equal probability of being allocated to each arm remaining in the trial including control. Both randomization algorithms began interim analyses after 100 patient-outcomes were available and repeated them every 14 days thereafter.

The response-adaptive randomization algorithm used for these simulations was a mixed patient-benefit and power-oriented approach analogous to that used in the REMAP trial and algorithms shown to have better performance among adaptive randomization algorithms.^2,23^ For probability *ρ*_*g*_ of allocation to treatment group *g*, probability *O(g)* that intervention *g* is optimal, and number of patients *n*_*g*_ allocated to group *g*:

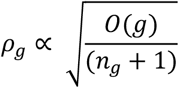

The algorithm additionally preserved power by stipulating that if any *ρ*_*g*_ < 0.10, it is increased and all *ρ*_*g*_ are renormalized such that all allocation probabilities are at least 0.10.

### Statistical treatment of time-varying outcomes

Analyses of trials using complete randomization can adjust for time to improve precision.^48^ For trials using RAR, adjustment for time can also reduce bias ^32,49^ but may not be able to accommodate the extreme time-trend shifts seen in pandemic epidemiology. The simulations compared trials with and without adjustment for time in interim and final analyses by dividing the timeline into 14-day “blocks” and analyzing time block as a categorical variable. Smaller time blocks and lower enrollment rates were investigated as sensitivity analyses to explore the performance of time trend adjustment with fewer patients in each time block.

### Stopping rules

The simulated trials used stopping rules analogous to those in REMAP and Thall et al.^2,10,11,21^ An intervention was declared superior if the probability it was the optimal intervention was greater than 0.99, and inferior if the same probability was less than 0.01. The region of practical equivalence ranged from an odds ratio of 1/1.2 to 1.2 and an intervention was declared equivalent if the probability of the odds ratio (relative to control) falling in that region was 0.90 or greater. Arms were dropped when determined to be inferior or equivalent. The trial stopped at 365 days, if an intervention was superior, or if all interventions were either inferior or equivalent.

### Estimand

The estimand for these simulations was the log-odds ratio associated with treatment.^33^ The odds ratio was assumed constant across varying baseline risk within each model.

### Methods for trial analysis

Interim and final analyses used Bayesian logistic regression to calculate the posterior distribution of the log-odds of mortality associated with each intervention.^50,51^ Models including adjustment for time incorporated time-block of enrollment as a categorical variable. A case with no interim analyses was considered as a sensitivity analysis.

### Performance measures

Performance measures were calculated for each factorial combination of the simulation characteristics. Results were also calculated according to the subgroup of whether or not the trial reached a conclusion, defined as either a single intervention being declared superior or all interventions being declared equivalent or inferior according to the stopping rules.

### Bias

The primary outcome for this simulation study was bias in the estimate of the odds ratio. This was calculated as the relative (percentage) difference between the mean estimate across simulations and the true value. We calculated Monte Carlo standard error and the root mean-squared error for the log-odds ratio.

### Operating characteristics

True negative rate was calculated as the proportion of simulated trials where an intervention with OR ≥ 1 did not meet the criterion for superiority. True positive rate was calculated as the proportion of trials that declared an effective intervention to be superior. Practical efficiency characteristics included the proportion of trials that reached a conclusion, the duration among trials that reached a conclusion, and the sample size enrolled in each arm.

### Ethical import

The ethical import of the trials was assessed by the proportion of patients randomized to a superior intervention where one existed, the time to dropping inferior or equivalent arms, and the average 28-day mortality of participants in the trial.

### Computational details

All simulations were coded in R and Stan using RStudio and several supporting packages.^52–58^ Existing software does not accommodate the time-adjustment aspect of these analyses.^59^ The Niagara computer cluster run by Compute Canada was used to run the simulations.^60,61^ Each Bayesian regression was run in 2 chains for 3000 iterations with 1000 iterations warmup. The number of simulated trials was chosen to be 1000 to keep the Monte Carlo standard error for the log-odds of bias approximately below 0.005, corresponding to a 95% confidence interval for odds ratio of bias 0.99 to 1.01.^33^ Chains and r-hat values were inspected in test cases to ensure convergence.^50^ All R code is available in the supporting information.

## Results

We simulated 1000 trials for each of 24 factorial scenarios. This required 3.5 hours of computation across 4 nodes on the cluster each using 80 cores (1120 core-hours of computation).

### Bias

Relative bias ranged from 0.3% to 11%, with variation according to scenario across the simulations (Table 1, Table E1). Across all scenarios with time-adjusted analyses, bias was less than 15% of the size of the average 95% credible interval for the mean odds ratio. In the null scenario the odds ratios were biased towards harm (OR > 1), and bias was largest (11%) in the single surge scenario using complete randomization without adjustment for time. Results were similar for interventions with OR set to 1.0 in the nugget scenario (Table E1), where complete randomization was associated with greater bias than adaptive randomization.

**Table 1:** Bias for equivalent and effective interventions

| Covid Scenario | Intervention Scenario<sup>a</sup> | Time adjustment | Algorithm | Mean posterior odds ratio of treatment<sup>b</sup> | Relative bias (%) | Bias (log-odds) | Monte Carlo SE<sup>c</sup> | RMSE<sup>d</sup> |
| --- | --- | --- | --- | --- | --- | --- | --- | --- |
| No surge | Null (OR = 1) | No | Complete | 1.04 (0.83 to 1.33) | 3.9 | 0.0389 | 0.0044 | 0.145 |
|  |  |  | Adaptive | 1.00 (0.82 to 1.25) | 0.4 | 0.0035 | 0.0042 | 0.134 |
|  |  | Yes | Complete | 1.06 (0.83 to 1.43) | 6.5 | 0.0646 | 0.0054 | 0.181 |
|  |  |  | Adaptive | 1.03 (0.83 to 1.32) | 2.7 | 0.0267 | 0.0053 | 0.171 |
|  | Nugget (OR = 0.75) | No | Complete | 0.74 (0.6 to 0.93) | 1.9 | 0.0187 | 0.0047 | 0.150 |
|  |  |  | Adaptive | 0.73 (0.59 to 0.92) | 3.2 | 0.0315 | 0.0048 | 0.154 |
|  |  | Yes | Complete | 0.75 (0.61 to 0.95) | 0.3 | 0.0032 | 0.0049 | 0.153 |
|  |  |  | Adaptive | 0.72 (0.58 to 0.92) | 4.3 | 0.0434 | 0.0047 | 0.156 |
| Single surge | Null (OR = 1) | No | Complete | 1.11 (0.91 to 1.44) | 11.4 | 0.1142 | 0.0069 | 0.244 |
|  |  |  | Adaptive | 1.04 (0.87 to 1.36) | 4.5 | 0.0445 | 0.0086 | 0.274 |
|  |  | Yes | Complete | 1.04 (0.84 to 1.32) | 3.8 | 0.0384 | 0.0043 | 0.142 |
|  |  |  | Adaptive | 1.02 (0.83 to 1.28) | 2.0 | 0.0195 | 0.0045 | 0.142 |
|  | Nugget (OR = 0.75) | No | Complete | 0.74 (0.61 to 0.94) | 0.9 | 0.0090 | 0.0051 | 0.162 |
|  |  |  | Adaptive | 0.67 (0.54 to 0.87) | 10.4 | 0.1043 | 0.0065 | 0.233 |
|  |  | Yes | Complete | 0.74 (0.61 to 0.92) | 1.4 | 0.0136 | 0.0045 | 0.142 |
|  |  |  | Adaptive | 0.72 (0.58 to 0.91) | 4.0 | 0.0399 | 0.0042 | 0.139 |
| Cyclic surges | Null (OR = 1) | No | Complete | 1.06 (0.87 to 1.34) | 6.2 | 0.0618 | 0.0051 | 0.172 |
|  |  |  | Adaptive | 1.03 (0.85 to 1.3) | 2.6 | 0.0259 | 0.0060 | 0.192 |
|  |  | Yes | Complete | 1.05 (0.84 to 1.34) | 4.7 | 0.0472 | 0.0045 | 0.149 |
|  |  |  | Adaptive | 1.02 (0.84 to 1.26) | 1.8 | 0.0177 | 0.0045 | 0.142 |
|  | Nugget (OR = 0.75) | No | Complete | 0.75 (0.62 to 0.92) | 0.6 | 0.0063 | 0.0044 | 0.138 |
|  |  |  | Adaptive | 0.7 (0.57 to 0.89) | 6.9 | 0.0691 | 0.0060 | 0.203 |
|  |  | Yes | Complete | 0.74 (0.61 to 0.91) | 1.9 | 0.0194 | 0.0040 | 0.129 |
|  |  |  | Adaptive | 0.72 (0.59 to 0.91) | 3.6 | 0.0363 | 0.0038 | 0.126 |

a: Null refers to all interventions having odds ratio 1; Nugget refers to intervention 2 having odds ratio 0.75 and all others 1.
b: The point estimate is the odds ratio associated with the mean posterior log-odds of treatment. In parentheses are the odds ratios associated with the mean lower and upper bounds of the 95% credible interval around the posterior log odds of treatment. This gives a sense of the average outcome estimate and 95% credible interval across simulated trials.
c: SE = standard error, in log-odds units
d: RMSE = root mean standard error, in log-odds units

In the nugget scenario, where one intervention had odds ratio 0.75, odds ratios generally overestimated benefit with the lowest odds ratio seen in the single surge nugget scenario using adaptive randomization without adjustment for time trends (OR estimate 0.69, relative bias 10.4%). Bias in this scenario decreased (OR 0.73, relative bias 4.0%) with the use of adjustment for time (Figure 2). Throughout interventions with odds ratio of 0.75, complete as opposed to adaptive randomization gave less biased estimates of the odds ratio.

**Figure 2:**
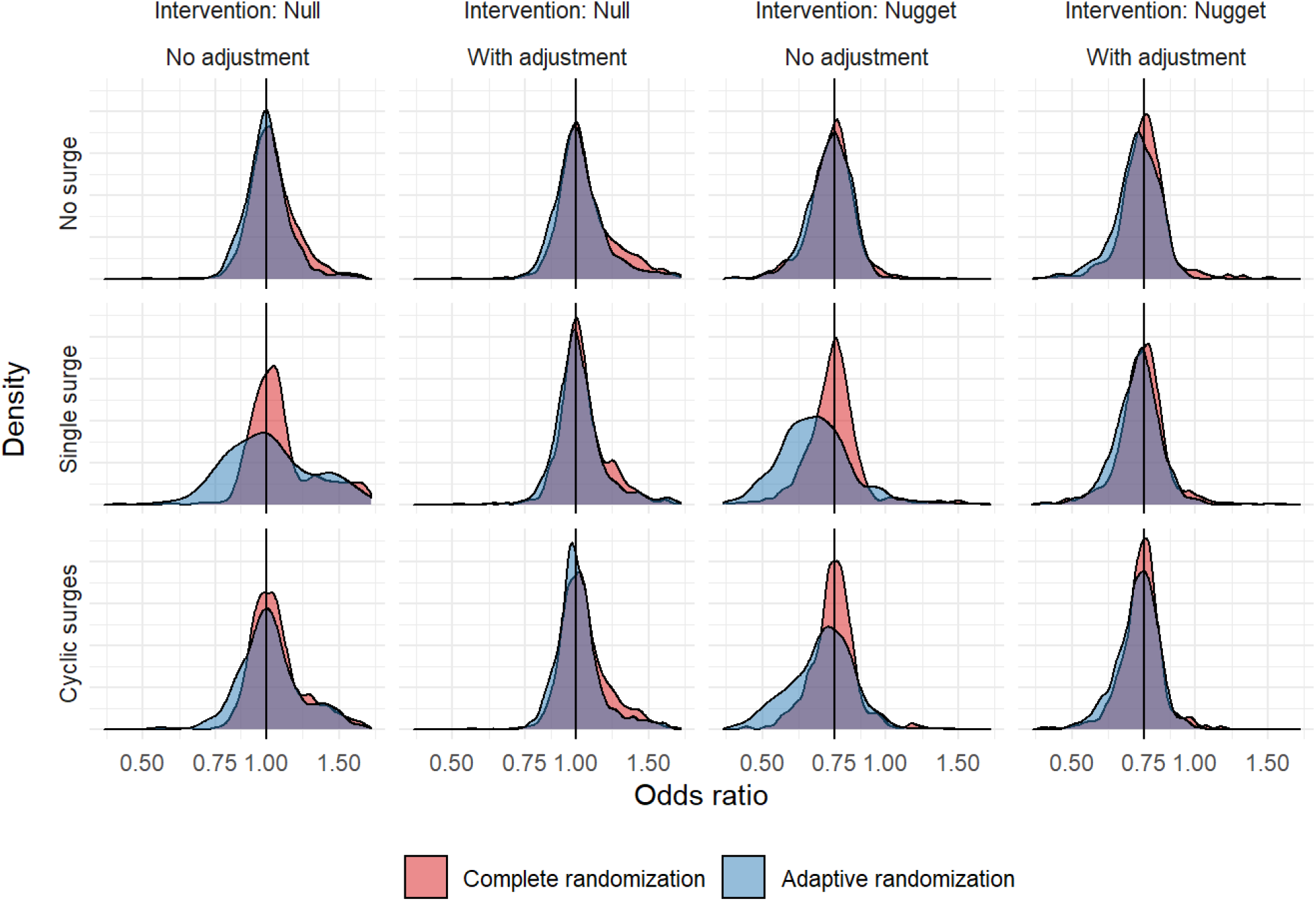
Odds ratio densities for Intervention 2 by COVID scenario. This figure depicts the distribution of 1000 posterior mean odds ratios for intervention 2 according to simulation scenario, including COVID epidemiology (rows: no surge, single surge, or cyclic surges), intervention scenario (columns: null, nugget), adjustment for time trends (columns: no adjustment, with adjustment), and randomization algorithm (colour). A black vertical line marks the set odds ratio for intervention 2 in either the “null” (OR 1) or “nugget” (OR 0.75) intervention scenarios. The improvement in bias with adjustment for time trends is most evident for adaptive randomization in the single surge scenario.

### Operating characteristics

The proportion of trials that appropriately concluded equivalent interventions were not superior was high (lowest proportion 0.958, Table 2). After incorporating adjustment for time, the proportion of trials using adaptive randomization that appropriately concluded equivalent interventions were not superior was 0.99 or higher in all scenarios (Table 2, Table E2). The proportion of trials where an ineffective intervention was declared optimal was low across null scenarios.

**Table 2:** Trial conclusions and rejection for equivalent and effective interventions

| Covid Scenario | Intervention Scenario | Time adjustment | Algorithm | Trials that reached a conclusion | Type of conclusion |  | Type of non-superiority |  |  | Monte Carlo SE for Rejection |
| --- | --- | --- | --- | --- | --- | --- | --- | --- | --- | --- |
|  |  |  |  |  | Superiority | Rejection<sup>a</sup> | Equivalence | Inferiority | Continue |  |
| No surge | Null (OR = 1) | No | Complete | 0.588 | 0.003 | 0.997 | 0.476 | 0.333 | 0.188 | 0.002 |
|  |  |  | Adaptive | 0.453 | 0.007 | 0.993 | 0.441 | 0.247 | 0.305 | 0.003 |
|  |  | Yes | Complete | 0.611 | 0.008 | 0.992 | 0.467 | 0.342 | 0.183 | 0.003 |
|  |  |  | Adaptive | 0.387 | 0.008 | 0.992 | 0.372 | 0.284 | 0.336 | 0.003 |
|  | Nugget (OR = 0.75) | No | Complete | 0.486 | 0.428 | 0.572 | 0.014 | 0.054 | 0.504 | 0.016 |
|  |  |  | Adaptive | 0.616 | 0.571 | 0.429 | 0.016 | 0.045 | 0.368 | 0.016 |
|  |  | Yes | Complete | 0.443 | 0.380 | 0.620 | 0.013 | 0.057 | 0.550 | 0.015 |
|  |  |  | Adaptive | 0.591 | 0.553 | 0.447 | 0.006 | 0.041 | 0.400 | 0.016 |
| Single surge | Null (OR = 1) | No | Complete | 0.710 | 0.006 | 0.994 | 0.513 | 0.344 | 0.137 | 0.002 |
|  |  |  | Adaptive | 0.505 | 0.042 | 0.958 | 0.202 | 0.486 | 0.270 | 0.006 |
|  |  | Yes | Complete | 0.696 | 0.005 | 0.995 | 0.565 | 0.286 | 0.144 | 0.002 |
|  |  |  | Adaptive | 0.496 | 0.002 | 0.998 | 0.461 | 0.262 | 0.275 | 0.001 |
|  | Nugget (OR = 0.75) | No | Complete | 0.793 | 0.717 | 0.283 | 0.016 | 0.065 | 0.202 | 0.014 |
|  |  |  | Adaptive | 0.861 | 0.770 | 0.230 | 0.042 | 0.060 | 0.128 | 0.013 |
|  |  | Yes | Complete | 0.535 | 0.466 | 0.534 | 0.025 | 0.051 | 0.458 | 0.016 |
|  |  |  | Adaptive | 0.675 | 0.616 | 0.384 | 0.018 | 0.054 | 0.312 | 0.015 |
| Cyclic surges | Null (OR = 1) | No | Complete | 0.834 | 0.003 | 0.997 | 0.554 | 0.364 | 0.079 | 0.002 |
|  |  |  | Adaptive | 0.735 | 0.024 | 0.976 | 0.404 | 0.438 | 0.134 | 0.005 |
|  |  | Yes | Complete | 0.802 | 0.004 | 0.996 | 0.577 | 0.328 | 0.091 | 0.002 |
|  |  |  | Adaptive | 0.654 | 0.005 | 0.995 | 0.557 | 0.258 | 0.180 | 0.002 |
|  | Nugget (OR = 0.75) | No | Complete | 0.761 | 0.675 | 0.325 | 0.034 | 0.056 | 0.235 | 0.015 |
|  |  |  | Adaptive | 0.852 | 0.764 | 0.236 | 0.049 | 0.045 | 0.142 | 0.013 |
|  |  | Yes | Complete | 0.584 | 0.527 | 0.473 | 0.013 | 0.046 | 0.414 | 0.016 |
|  |  |  | Adaptive | 0.768 | 0.713 | 0.287 | 0.014 | 0.044 | 0.229 | 0.014 |

a. Rejection encompasses any conclusion other than superiority (continue, equivalence, or inferiority).

The proportion of trials that reached a conclusion varied according to pandemic epidemiology from a low of 0.39 (null scenario, no surge, adaptive randomization, adjustment for time trends) to a high of 0.86 (nugget scenario, single surge, adaptive randomization, no adjustment for time trends).

The frequency with which superior interventions were identified as such was not high in any scenario. For interventions with odds ratio 0.75, the lowest probability of being declared superior (0.38) was seen in simulations with no surge, complete randomization, and adjustment for time trends. By contrast, the highest probability of being declared superior (0.77) was seen in simulations with a single surge, adaptive randomization, and no adjustment for time trends (Table 2, Figure E1). Trials using adaptive as opposed to complete randomization in the nugget scenario yielded a higher probability of identifying a superior intervention (Figure 3).

**Figure 3:**
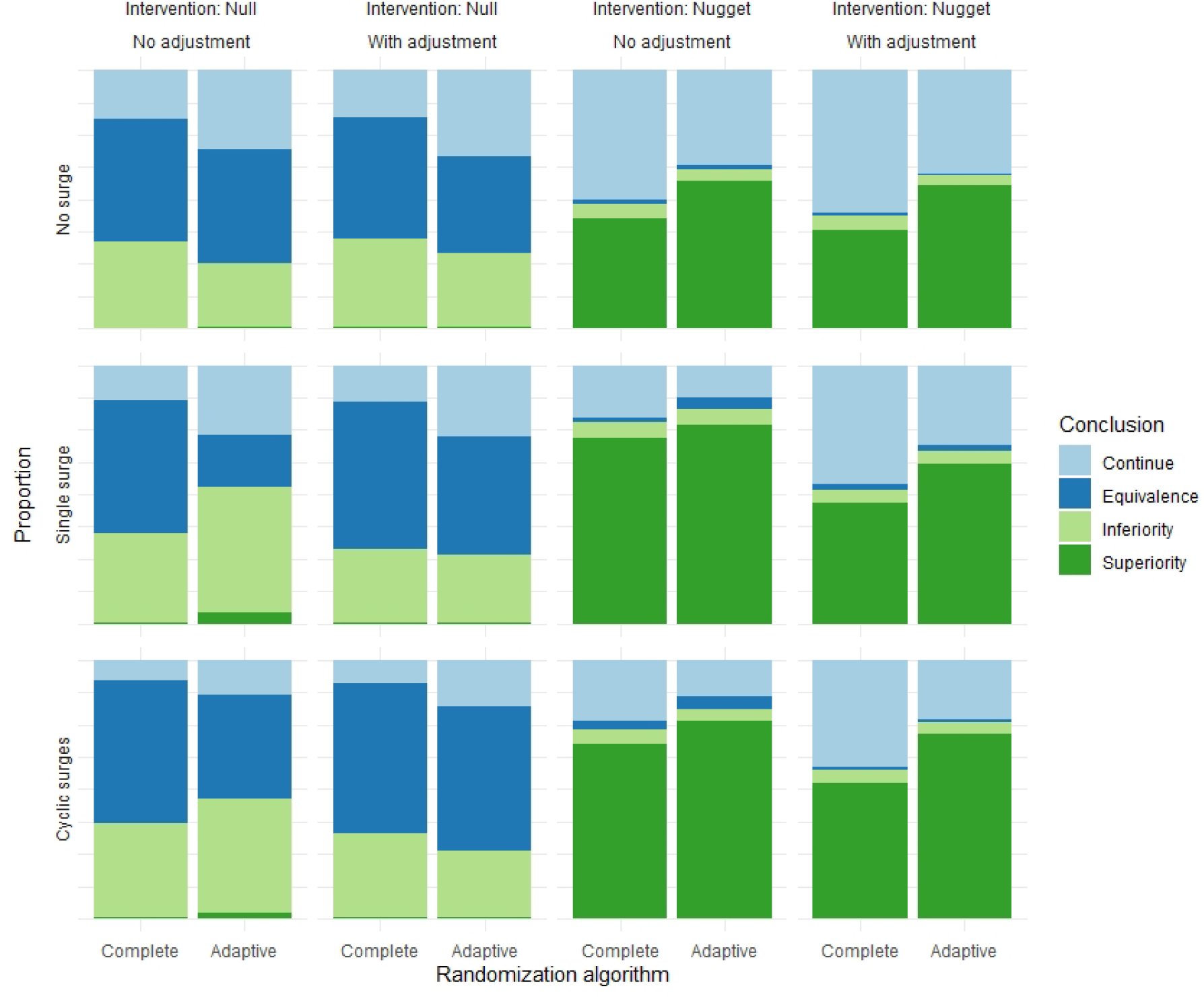
Trial conclusions for equivalent and effective interventions. This figure depicts the proportion of trials that concluded continue, equivalence, inferiority, or superiority for intervention 2 according to simulation scenario, including COVID epidemiology (rows: no surge, single surge, or cyclic surges), intervention scenario (columns: null, nugget), adjustment for time trends (columns: no adjustment, with adjustment), and randomization algorithm (colour). Adaptive as opposed to complete randomization results in a higher proportion of trials concluding that intervention 2 is superior in the nugget scenario.

Among trials that did not reach conclusions according to the stopping criteria, the probability that an effective intervention was the optimal arm was still high. In those trials, the highest mean posterior probability of an effective intervention being optimal (0.945) was seen in the cyclic surge scenario with adaptive randomization and adjustment for time trends (Table E3).

Sample size was generally large (Table 3). Because most trials were limited by the 365-day time-limit, sample sizes were similar across intervention scenarios.

**Table 3:** Outcomes of trial for equivalent and effective interventions

| Covid Scenario | Intervention Scenario<sup>1</sup> | Time adjustment | Algorithm | Probability of being optimal<sup>a</sup> | Probability of being equivalent | Sample size: Control arm | Sample size: Intervention 2 | Sample size: Intervention 3 | Sample size: Total | Mortality rate |
| --- | --- | --- | --- | --- | --- | --- | --- | --- | --- | --- |
| No surge | Null (OR = 1) | No | Complete | 0.236 | 0.742 | 1578 | 1114 | 1095 | 4908 | 0.200 |
|  |  |  | Adaptive | 0.256 | 0.765 | 1334 | 1266 | 1288 | 5130 | 0.200 |
|  |  | Yes | Complete | 0.241 | 0.697 | 1575 | 1070 | 1133 | 4880 | 0.200 |
|  |  |  | Adaptive | 0.243 | 0.724 | 1357 | 1243 | 1272 | 5163 | 0.200 |
|  | Nugget (OR = 0.75) | No | Complete | 0.903 | 0.194 | 1599 | 1538 | 686 | 4512 | 0.186 |
|  |  |  | Adaptive | 0.951 | 0.202 | 835 | 2200 | 702 | 4438 | 0.179 |
|  |  | Yes | Complete | 0.898 | 0.199 | 1652 | 1580 | 703 | 4640 | 0.186 |
|  |  |  | Adaptive | 0.951 | 0.190 | 831 | 2172 | 700 | 4396 | 0.179 |
| Single surge | Null (OR = 1) | No | Complete | 0.226 | 0.659 | 1557 | 1062 | 1061 | 4786 | 0.225 |
|  |  |  | Adaptive | 0.259 | 0.490 | 1222 | 1215 | 1195 | 4807 | 0.226 |
|  |  | Yes | Complete | 0.250 | 0.756 | 1570 | 1138 | 1069 | 4880 | 0.224 |
|  |  |  | Adaptive | 0.248 | 0.763 | 1397 | 1282 | 1292 | 5264 | 0.222 |
|  | Nugget (OR = 0.75) | No | Complete | 0.932 | 0.199 | 1379 | 1317 | 668 | 4026 | 0.218 |
|  |  |  | Adaptive | 0.909 | 0.141 | 726 | 1549 | 647 | 3573 | 0.218 |
|  |  | Yes | Complete | 0.910 | 0.194 | 1626 | 1567 | 706 | 4550 | 0.213 |
|  |  |  | Adaptive | 0.955 | 0.183 | 812 | 2195 | 677 | 4363 | 0.208 |
| Cyclic surges | Null (OR = 1) | No | Complete | 0.239 | 0.722 | 1561 | 1066 | 1046 | 4777 | 0.258 |
|  |  |  | Adaptive | 0.244 | 0.661 | 1199 | 1160 | 1165 | 4712 | 0.259 |
|  |  | Yes | Complete | 0.241 | 0.750 | 1612 | 1103 | 1127 | 4963 | 0.258 |
|  |  |  | Adaptive | 0.249 | 0.780 | 1447 | 1303 | 1334 | 5406 | 0.257 |
|  | Nugget (OR = 0.75) | No | Complete | 0.921 | 0.201 | 1520 | 1459 | 682 | 4350 | 0.242 |
|  |  |  | Adaptive | 0.929 | 0.183 | 771 | 1814 | 684 | 3941 | 0.236 |
|  |  | Yes | Complete | 0.924 | 0.163 | 1705 | 1654 | 674 | 4702 | 0.241 |
|  |  |  | Adaptive | 0.962 | 0.175 | 826 | 2253 | 678 | 4435 | 0.235 |

a: These results are averaged across all 1000 trials for each scenario. For example, the column “Probability of being optimal” reports the mean probability that intervention 2 is optimal across all 1000 trials in each scenario.

### Ethical import

In the nugget scenario, trials using adaptive randomization consistently randomized about twice as many patients to the superior intervention arm (Table 3). The average risks of 28-day mortality were lower in trials using adaptive randomization, although absolute differences were small (Table 3, Figure E2).

Equivalent or harmful interventions were generally dropped earlier from trials using complete as opposed to adaptive randomization (Figure E3). By contrast, there was minimal difference in time to recognition of a superior intervention.

A large number of null interventions (OR 1) had “Inferiority” conclusions. Across all factorial scenarios, in 95% of trials where intervention 3 (OR = 1) was deemed inferior the posterior probability of equivalence with respect to control was at least 0.78.

## Discussion

This simulation study of COVID-19 found that response-adaptive randomization can generate biased estimates of efficacy amidst surges of COVID-19, and that adjustment for time eliminates this bias. Simulations used realistic trial design and parameters. The bias was generally in the direction of overestimating the efficacy of effective interventions. Relative to complete randomization, response-adaptive randomization with time-adjustment increased the number of patients allocated to effective interventions, increased the probability of identifying effective interventions, and did not increase the probability of concluding that equivalent interventions were effective. However, it delayed the removal of ineffective interventions.

Adjustment for time trends during pandemic surges attenuated bias for both adaptive and complete randomization approaches, but it slightly increased bias if there was no surge. However, the slight increase in bias in the case of no surge is outweighed by the alleviation of large amounts of bias in the cases with surge, so where numerically feasible, adjustment for time trends should be the standard-of-design for Bayesian trials using either response-adaptive randomization or complete randomization with frequent interim analyses.

The simulations revealed important consequences of using complete versus response-adaptive randomization. Trials using complete randomization identified ineffective interventions earlier than trials using response-adaptive randomization. This occurred because in response-adaptive randomization fewer patients are allocated to interventions that are measured to be ineffective, so more time is required to recruit enough patients to definitively remove those interventions from the trial. The proportion of all COVID-19 patients enrolled in trials is low, making this an important consideration in trial design. Announcement of the RECOVERY trial hydroxychloroquine results on June 5, 2020 ^62^, for example, allowed other trials such as the SOLIDARITY trial to stop allocating patients to hydroxychloroquine, and hopefully reduced the inappropriate use of hydroxychloroquine worldwide. More subtle potential benefits include redirection of resources to other avenues for reducing COVID-19 morbidity and mortality, such as additional experimental agents, methods of contact tracing and isolation, or public health campaigns to promote universal masking.

Trials using response-adaptive randomization were more likely to identify effective interventions than trials using complete randomization. Even among trials that did not reach a definite conclusion by 365 days, effective interventions studied with adaptive randomization had a higher probability of being the optimal intervention compared to those studied with complete randomization. Historically, the high proportion of trials with indeterminate or negative results has been a source of frustration for clinicians and trialists.^63^ Although adaptive randomization does not increase the chance that an intervention is effective, it does increase the chance that an effective intervention is appropriately recognized.

Response-adaptive randomization also led to more patients being allocated to an effective intervention, when one was present. This could help increase patient enrollment by satisfying an individual patient’s desire to have improved outcomes through participation in research.^7^ This may also encourage clinicians to participate in the trial by reducing the tension between desire to give an individual patient a potentially optimal therapy and the need for randomized evaluation where equipoise exists. However, increased complexity of the consent process and trial conduct may offset these benefits.^64^

On other aspects of trial performance, the two randomization algorithms were largely similar. Despite randomizing many more patients to effective interventions, trials using response-adaptive randomization did not show a clinically significant improvement in average patient outcomes, partially due to the small difference (20% versus 15.8%) in mortality rates outside of surge conditions. After incorporating adjustment for time trends, there was little difference in true negative rates.

This investigation has several limitations. First, response-adaptive randomization algorithms are not monolithic and different implementations of RAR may lead to different conclusions.^32,65^ Investigation of all possible RAR design choices is infeasible, so instead we modeled our algorithm after a large ongoing trial of critically ill patients with COVID-19. Second, our implementation of COVID epidemiology may not be a nuanced enough representation of the actual dynamics of mortality or enrollment rates during a surge in cases. Third, the validity of these results may depend on having enough patients in each “time block” such that categorical adjustment for time trends yields an accurate model. Fourth, the simulations modeled a multicenter trial with a surge taking place at the same time in every center, a pattern not seen with COVID-19 surges so far. If surges occurred throughout a trial, and at different times at different sites, the net effect would be regression to the mean event rate, so the issues with RAR could be less pronounced than in the scenarios modeled here. Finally, while the code for replication of this study is easily available, the computational demands of the simulations may limit efforts to plan Bayesian adaptive trials with adjustment for time trends.^36,66^

The decision to incorporate adaptive versus complete randomization into a trial of a therapy for COVID-19 also depends on factors beyond those captured in these simulations. Adaptive randomization introduces the possibility of clinicians inferring interim results due to detectable changes in treatment allocation as the trial progresses. Using adaptive randomization requires an added level of organization and communication across sites that may outweigh any statistical or ethical benefits amidst the chaos of a pandemic. If adaptive randomization is used with adjustment for time, meta-analyses of therapies would require individual patient data. Finally, research concerning COVID-19 is reaching a much wider audience than almost any medical research previously undertaken, potentially increasing the requirement for simple and transparent statistical analyses as opposed to the more complex analyses required for adaptive randomization. Optimal trial design requires consideration of both quantitative and qualitative aspects of design elements.

## Conclusion

In trials of therapies for COVID-19 during surges that may be associated with increased mortality rates, the use of response-adaptive randomization without adjustment for time estimates results in biased estimates of efficacy. However, this bias is eliminated with adjustment for time and response-adaptive as opposed to complete randomization has a higher probability of identifying effective interventions. Response-adaptive randomization with time adjustment is an appealing option for pandemic trials.

## Data Availability

All data produced in the present study are available upon reasonable request to the authors

## Acknowledgements

We thank the following for helpful comments: Scott Berry.

