## Supplemental appendix for "COVID-19 pandemic surges can induce bias in trials using response adaptive randomization: A simulation study"

#### Supplement

Yarnell, Christopher J MD FRCP(C); ORCID 0000-0001-5657-9398

Fowler, Robert A MDCM MS(Epi) FRCP(C)

Sung, Lillian

Tomlinson, George PhD

2020-10-15

### Additional methods background

#### Pandemic epidemiology

Pandemic epidemiology was represented through both the 28-day mortality rate and the enrollment rate. If a surge of COVID-19 pneumonia cases overburdens acute care capacity at centers in a trial, it may cause shortages with respect to medications, equipment, physical locations for care, infection control, and clinical staff with corresponding increases in both enrollment and 28-day mortality rates during the surge. Surges of this nature have been observed in China [1], Italy [2], New York [3], Spain [4], and Brazil [5].

Information about the 28-day mortality rate for patients hospitalized due to COVID-19 pneumonia is accumulating and in flux. A recent meta-analysis of completed outcomes for patients admitted to intensive care units (ICUs) incorporating data from Asia, North America, and Europe, including 8,062 patients from the United Kingdom, showed an ICU mortality of 41.6%.[6,7] The preprint of the RECOVERY trial of dexamethasone use in hospitalized COVID-19 patients in the United Kingdom reported 28-day mortality on 4,321 patients allocated to usual care and observed a 24.6% mortality rate. A case series from New York with limited follow-up duration documented a 21% mortality rate among 2,634 patients either discharged or deceased.[3] Based on these rates, we selected a 28-day mortality rate of 20% as the base case. Observing that narrative reports of the experience in Italy, Spain, and New York describe critical care being provided throughout the hospital, we estimate that during a surge the mortality of hospitalized patients approximates the mortality rate of patients admitted to the ICU and we set the surge case fatality rate to 40%.

The number of cases per day requiring hospitalization is commonly reported in COVID-19 epidemiology.[3,8,9] The relationship between new hospitalized cases per day and enrollment rates is less certain. The REMAP trial has enrolled approximately 100 patients every 2 weeks during the spring of 2020, with few sites ready to enroll during the first wave of COVID-19, while the RECOVERY trial enrolled 6,425 patients over approximately 90 days amounting to about 70 patients per day.[10] We compromised on a rate of 15 patients per day, approximating an international multicenter trial. Based on an uncertain ability to ramp up enrollment proportionally to the influx of new cases during a pandemic surge, we chose a conservative increase in mean enrolment from 15 to 25 patients per day.

The duration of each surge was set to 21 days, based on the large dataset from the United Kingdom.[7] The first surge, when present, began 30 days after the start of the trial. The cyclic surge scenario used three surges (spring, fall, winter) of 21 days duration starting on day 30, 150, and 270 of the trial, approximately similar to the 1918 influenza pandemic.[11] The base case had no surge, and the two alternative cases had a single surge and cyclic surges. Single surge cases with later surge timing (days 150 and 270) were investigated in sensitivity analyses.

#### Number and efficacy of intervention arms

The trials evaluated three intervention arms and one control arm, mimicking large trials of COVID-19 evaluating multiple arms: the RECOVERY trial [10], the REMAP trial [12], and the global SOLIDARITY trial coordinated by the World Health Organization (WHO) [13]. Review of the International Clinical Trials Registry Platform organized by the WHO shows a total of 1,427 interventional trials of which 203 (14%) are multi-arm.[14] Multi-arm trials may be preferable to single-arm trials in the setting of a pandemic because the efficiency gains due to shared control patients and built-in comparative effectiveness information may increase the speed of discovery while also allowing for simultaneous testing of multiple incompletely studied interventions for a new disease.[15,16]

Preliminary results from COVID-19 trials, as well as past results from trials in the acute respiratory distress syndrome, gave some indication as to plausible odds ratios for interventions. The most common scenario in COVID-19 trials is likely to be the null scenario, where all interventions have an odds ratio near 1.[17,18] In the scenario where an intervention shows benefit, preliminary data concerning remdesivir [19] and dexamethasone [10] suggest a plausible range of odds ratios. Remdesivir was associated with a hazard ratio for death at 14 days of 0.70.[19] In the RECOVERY trial comparing patients who did and did not receive dexamethasone, the odds ratio for 28-day mortality was 0.60 in patients requiring invasive mechanical ventilation and 0.82 in patients requiring oxygen therapy but not invasive mechanical ventilation.[10] For the "nugget" scenario where one intervention is effective and the remainder have odds ratio set to 1.0, we set the effective intervention to have an odds ratio of 0.75. The control arm was always intervention 1 and if there was an effective intervention, it was always intervention 2. As sensitivity analyses, we also conducted simulations in the scenario where a single intervention was harmful with an odds ratio of 1.33 and where there was one effective intervention, one harmful intervention, and one equivalent intervention.[20]

#### Randomization algorithms

The base case for randomization was complete randomization where throughout the trial patients have equal probability of being allocated to control and each intervention arm remaining in the trial.

Response-adaptive randomization algorithms have multiple components: the number of patients enrolled before beginning to adapt, the frequency of adaptations or interim analyses, the equations that determine the allocation probabilities, and the method of allocation to the control group. [21–23] For this study, both randomization algorithms used the same minimum number of patients with outcomes available before conducting an interim analysis (100) and performed interim analyses every 14 days from that day. A case with no interim analyses was considered as a sensitivity analysis.

The equations that determine allocation probabilities vary according to the extent to which they are oriented towards patient benefit or towards statistical power.[24] The archetypal patient-benefit-oriented approach is known as Thompson Sampling where the probability of allocation to an intervention is a function of the probability that that arm is superior to all other interventions.[25] Thompson Sampling algorithms are vulnerable to bias due to time trends, and, due to randomness, are not guaranteed to allocate more patients to a superior intervention.[24,26] For example, if many patients assigned to a beneficial intervention early in a trial have a bad outcome, the adaptive randomization may subsequently assign fewer patients to that intervention. Efforts to attenuate the variance introduced by Thompson sampling generally shrink the distribution of allocation probabilities back towards complete randomization and increase the extent to which an equation is power-oriented.

The response-adaptive randomization algorithm used for these simulations was a mixed patient-benefit and power-oriented approach analogous to that used in the REMAP trial, to provide both pragmatic and academic insights, and based on the work of Wathen et al showing better performance among adaptive randomization algorithms using a similar approach.[12,27] For probability $\rho_g$ of allocation to treatment group $g$, probability $O(g)$ that intervention $g$ is optimal, and number of patients $n_g$ allocated to group $g$:

$$\rho_g \propto \sqrt{\frac{O(g)}{(n_g + 1)}}$$

Including the number of patients already allocated to that intervention in the denominator and taking the square root of the fraction increases the statistical power of adaptation over adaptation based on only $O(g)$. The algorithm additionally preserves power by stipulating that if any $\rho_g < 0.10$, it is increased and all $\rho_g$ are renormalized such that all allocation probabilities are at least 0.10.

#### Results for null interventions (OR = 1) in nugget and null scenarios

##### Table E1: Bias for Intervention 3

| Covid Scenario | Intervention Scenario[a] | Time trends modeled | Algorithm | Mean posterior odds ratio of treatment[b] | Bias (log-odds) | Monte Carlo SE[c] | RMSE[d] |
|---|---|---|---|---|---|---|---|
| No surge | Null | No | Complete | 1.04 (0.82 to 1.32) | 0.0300 | 0.0044 | 0.138 |
| | | | Adaptive | 1.01 (0.82 to 1.25) | -0.0004 | 0.0042 | 0.131 |
| | | Yes | Complete | 1.07 (0.83 to 1.39) | 0.0513 | 0.0054 | 0.164 |
| | | | Adaptive | 1.03 (0.83 to 1.29) | 0.0176 | 0.0053 | 0.150 |
| | Nugget | No | Complete | 1.04 (0.79 to 1.39) | 0.0314 | 0.0047 | 0.151 |
| | | | Adaptive | 1.04 (0.78 to 1.39) | 0.0256 | 0.0048 | 0.161 |
| | | Yes | Complete | 1.09 (0.8 to 1.5) | 0.0697 | 0.0049 | 0.186 |
| | | | Adaptive | 1.07 (0.79 to 1.46) | 0.0486 | 0.0047 | 0.192 |
| Single surge | Null | No | Complete | 1.15 (0.92 to 1.45) | 0.1112 | 0.0069 | 0.251 |
| | | | Adaptive | 1.09 (0.88 to 1.37) | 0.0530 | 0.0086 | 0.273 |
| | | Yes | Complete | 1.06 (0.84 to 1.36) | 0.0498 | 0.0043 | 0.161 |
| | | | Adaptive | 1.03 (0.83 to 1.27) | 0.0160 | 0.0045 | 0.141 |
| | Nugget | No | Complete | 1.22 (0.94 to 1.58) | 0.1698 | 0.0051 | 0.292 |
| | | | Adaptive | 1.1 (0.83 to 1.46) | 0.0591 | 0.0065 | 0.257 |
| | | Yes | Complete | 1.06 (0.8 to 1.41) | 0.0451 | 0.0045 | 0.161 |
| | | | Adaptive | 1.05 (0.79 to 1.41) | 0.0397 | 0.0042 | 0.167 |
| Cyclic surges | Null | No | Complete | 1.07 (0.87 to 1.33) | 0.0571 | 0.0051 | 0.166 |
| | | | Adaptive | 1.04 (0.84 to 1.29) | 0.0216 | 0.0060 | 0.183 |
| | | Yes | Complete | 1.05 (0.84 to 1.33) | 0.0424 | 0.0045 | 0.145 |
| | | | Adaptive | 1.03 (0.84 to 1.26) | 0.0159 | 0.0045 | 0.140 |
| | Nugget | No | Complete | 1.11 (0.86 to 1.42) | 0.0827 | 0.0044 | 0.201 |
| | | | Adaptive | 1.06 (0.81 to 1.39) | 0.0365 | 0.0060 | 0.205 |
| | | Yes | Complete | 1.06 (0.8 to 1.41) | 0.0461 | 0.0040 | 0.161 |
| | | | Adaptive | 1.05 (0.8 to 1.4) | 0.0368 | 0.0038 | 0.169 |

a: Null refers to all interventions having odds ratio 1; Nugget refers to intervention 2 having odds ratio 0.75 and all others 1.

b: The point estimate is the odds ratio associated with the mean posterior log-odds of treatment. In parentheses are the odds ratios associated with the mean lower and upper bounds of the 95% credible interval around the posterior log odds of treatment. This gives a sense of the average outcome estimate and 95% credible interval across simulated trials.

c: SE = standard error, in log-odds units

d: RMSE = root mean standard error, in log-odds units

#### Table E2: Trial conclusions and rejection for a null intervention (OR = 1) in nugget and null scenarios

| Covid Scenario | Intervention Scenario | Time adjustment | Algorithm | Trials that reached a conclusion | Type of conclusion | | Type of non-superiority | | | Monte Carlo SE for Rejection |
|---|---|---|---|---|---|---|---|---|---|---|
| | | | | | Superiority | Rejection[a] | Equivalence | Inferiority | Continue | |
| No surge | Null (OR = 1) | No | Complete | 0.994 | 0.006 | 0.456 | 0.329 | 0.209 | 0.002 | 0.994 |
| | | | Adaptive | 0.996 | 0.004 | 0.482 | 0.222 | 0.292 | 0.002 | 0.996 |
| | | Yes | Complete | 0.997 | 0.003 | 0.496 | 0.309 | 0.192 | 0.002 | 0.997 |
| | | | Adaptive | 0.996 | 0.004 | 0.389 | 0.255 | 0.352 | 0.002 | 0.996 |
| | Nugget (OR = 0.75) | No | Complete | 0.996 | 0.004 | 0.091 | 0.843 | 0.062 | 0.002 | 0.996 |
| | | | Adaptive | 1.000 | 0.000 | 0.037 | 0.883 | 0.080 | 0.000 | 1.000 |
| | | Yes | Complete | 0.997 | 0.003 | 0.103 | 0.844 | 0.050 | 0.002 | 0.997 |
| | | | Adaptive | 1.000 | 0.000 | 0.027 | 0.874 | 0.099 | 0.000 | 1.000 |
| Single surge | Null (OR = 1) | No | Complete | 0.990 | 0.010 | 0.509 | 0.350 | 0.131 | 0.003 | 0.990 |
| | | | Adaptive | 0.966 | 0.034 | 0.210 | 0.514 | 0.242 | 0.006 | 0.966 |
| | | Yes | Complete | 0.993 | 0.007 | 0.500 | 0.347 | 0.146 | 0.003 | 0.993 |
| | | | Adaptive | 0.992 | 0.008 | 0.470 | 0.257 | 0.265 | 0.003 | 0.992 |
| | Nugget (OR = 0.75) | No | Complete | 0.997 | 0.003 | 0.052 | 0.908 | 0.037 | 0.002 | 0.997 |
| | | | Adaptive | 0.997 | 0.003 | 0.030 | 0.922 | 0.045 | 0.002 | 0.997 |
| | | Yes | Complete | 0.994 | 0.006 | 0.102 | 0.847 | 0.045 | 0.002 | 0.994 |
| | | | Adaptive | 0.995 | 0.005 | 0.034 | 0.895 | 0.066 | 0.002 | 0.995 |
| Cyclic surges | Null (OR = 1) | No | Complete | 0.993 | 0.007 | 0.576 | 0.339 | 0.078 | 0.003 | 0.993 |
| | | | Adaptive | 0.986 | 0.014 | 0.415 | 0.427 | 0.144 | 0.004 | 0.986 |
| | | Yes | Complete | 0.995 | 0.005 | 0.582 | 0.311 | 0.102 | 0.002 | 0.995 |
| | | | Adaptive | 0.998 | 0.002 | 0.566 | 0.237 | 0.195 | 0.001 | 0.998 |
| | Nugget (OR = 0.75) | No | Complete | 0.998 | 0.002 | 0.068 | 0.907 | 0.023 | 0.001 | 0.998 |
| | | | Adaptive | 0.999 | 0.001 | 0.043 | 0.934 | 0.022 | 0.001 | 0.999 |
| | | Yes | Complete | 0.997 | 0.003 | 0.087 | 0.885 | 0.025 | 0.002 | 0.997 |
| | | | Adaptive | 0.999 | 0.001 | 0.042 | 0.926 | 0.031 | 0.001 | 0.999 |

#### Table E3: Outcomes of trials that did not reach conclusion by 365 days

| Covid Scenario | Intervention Scenario[1] | Time trends modeled | Algorithm | Probability of being optimal | Probability of being equivalent | Sample size: Control arm | Sample size: Intervention 2 | Sample size: Intervention 3 | Sample size: Total | Mortality rate |
|---|---|---|---|---|---|---|---|---|---|---|
| No surge | Null | No | Complete | 0.260 | 0.760 | 1650 | 1257 | 1295 | 5472 | 0.199 |
| | | | Adaptive | 0.279 | 0.741 | 1294 | 1421 | 1398 | 5469 | 0.200 |
| | | Yes | Complete | 0.278 | 0.738 | 1671 | 1284 | 1229 | 5472 | 0.196 |
| | | | Adaptive | 0.258 | 0.740 | 1329 | 1381 | 1357 | 5475 | 0.195 |
| | Nugget | No | Complete | 0.871 | 0.254 | 1998 | 1958 | 761 | 5475 | 0.185 |
| | | | Adaptive | 0.930 | 0.344 | 1029 | 2606 | 915 | 5465 | 0.180 |
| | | Yes | Complete | 0.885 | 0.260 | 2005 | 1983 | 731 | 5475 | 0.181 |
| | | | Adaptive | 0.922 | 0.310 | 1016 | 2591 | 947 | 5472 | 0.175 |
| Single surge | Null | No | Complete | 0.260 | 0.687 | 1738 | 1328 | 1325 | 5701 | 0.219 |
| | | | Adaptive | 0.275 | 0.482 | 1230 | 1467 | 1521 | 5690 | 0.219 |
| | | Yes | Complete | 0.269 | 0.733 | 1720 | 1305 | 1324 | 5693 | 0.198 |
| | | | Adaptive | 0.283 | 0.761 | 1361 | 1478 | 1452 | 5695 | 0.196 |
| | Nugget | No | Complete | 0.904 | 0.324 | 1969 | 1926 | 893 | 5696 | 0.204 |
| | | | Adaptive | 0.860 | 0.246 | 993 | 2317 | 1150 | 5691 | 0.201 |
| | | Yes | Complete | 0.898 | 0.232 | 2098 | 2071 | 791 | 5695 | 0.182 |
| | | | Adaptive | 0.930 | 0.336 | 1054 | 2780 | 929 | 5688 | 0.179 |
| Cyclic surges | Null | No | Complete | 0.291 | 0.740 | 1920 | 1428 | 1437 | 6130 | 0.254 |
| | | | Adaptive | 0.313 | 0.660 | 1089 | 1703 | 1615 | 6133 | 0.254 |
| | | Yes | Complete | 0.289 | 0.758 | 1946 | 1360 | 1422 | 6142 | 0.197 |
| | | | Adaptive | 0.293 | 0.747 | 1383 | 1608 | 1550 | 6136 | 0.197 |
| | Nugget | No | Complete | 0.891 | 0.265 | 2382 | 2354 | 696 | 6138 | 0.235 |
| | | | Adaptive | 0.906 | 0.314 | 1088 | 2791 | 1146 | 6139 | 0.232 |
| | | Yes | Complete | 0.900 | 0.208 | 2405 | 2398 | 667 | 6139 | 0.179 |
| | | | Adaptive | 0.945 | 0.320 | 1118 | 3068 | 961 | 6128 | 0.180 |

#### Additional Figures

##### Figure E1: Probability that equivalent and effective interventions are optimal

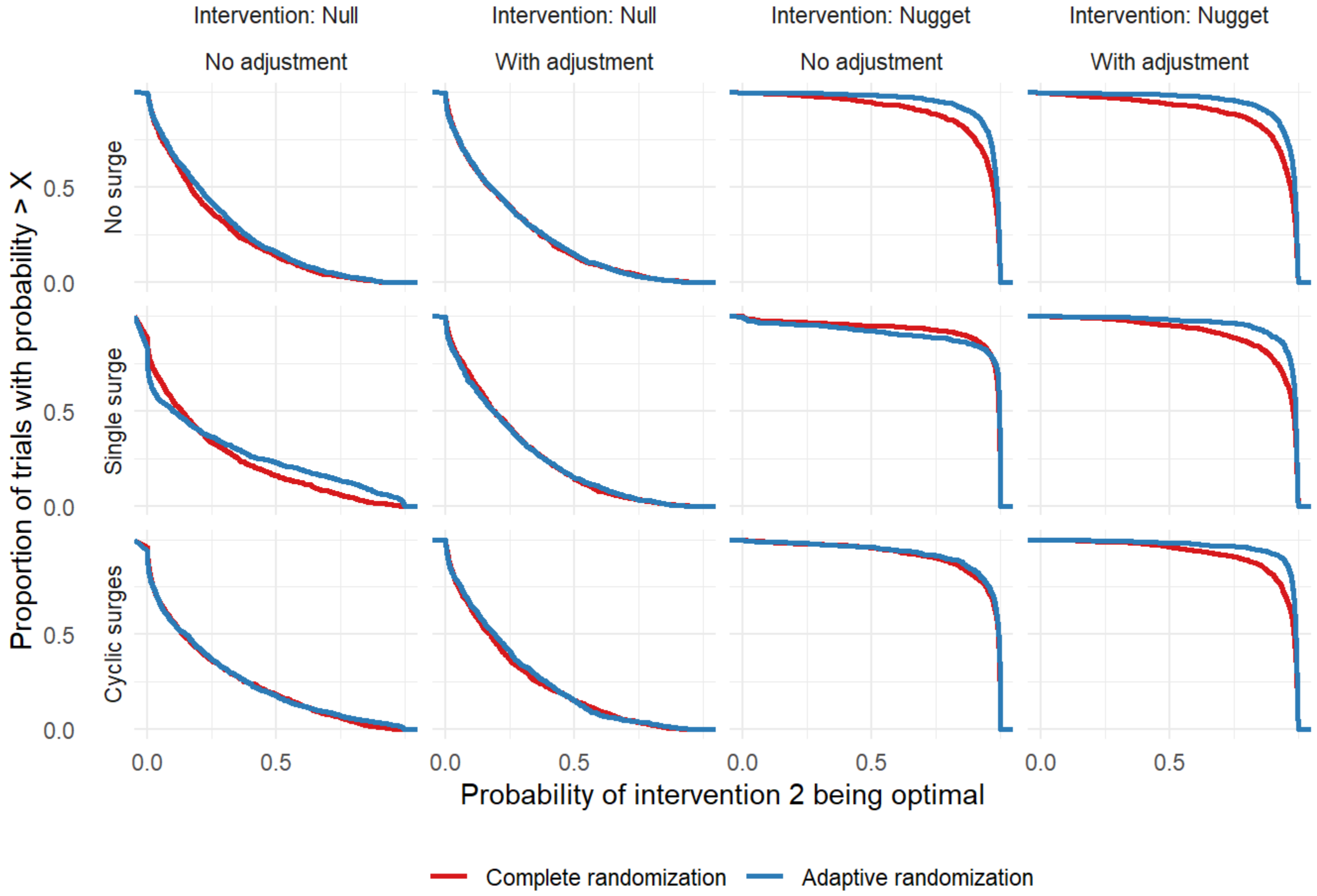

Figure E1 caption: This figure shows the proportions of trials finding intervention 2 to be optimal (y-axis) at different thresholds for declaring it to be optimal (x-axis). In this modeled trial, a threshold probability of 0.99 was used for superiority to be declared. The figure shows that in the null scenarios, fewer than 25% of trials return a probability > 0.50 that intervention 2 is optimal and almost no trials return a probability > 0.99 of intervention 2 being optimal. By contrast, almost all trials in the nugget scenario have a probability > 0.95 of intervention 2 being optimal. In all nugget scenarios with adjustment for time, more trials with adaptive randomization than with complete randomization exceed any given threshold.

#### Figure E2: Average outcome according to simulation scenario

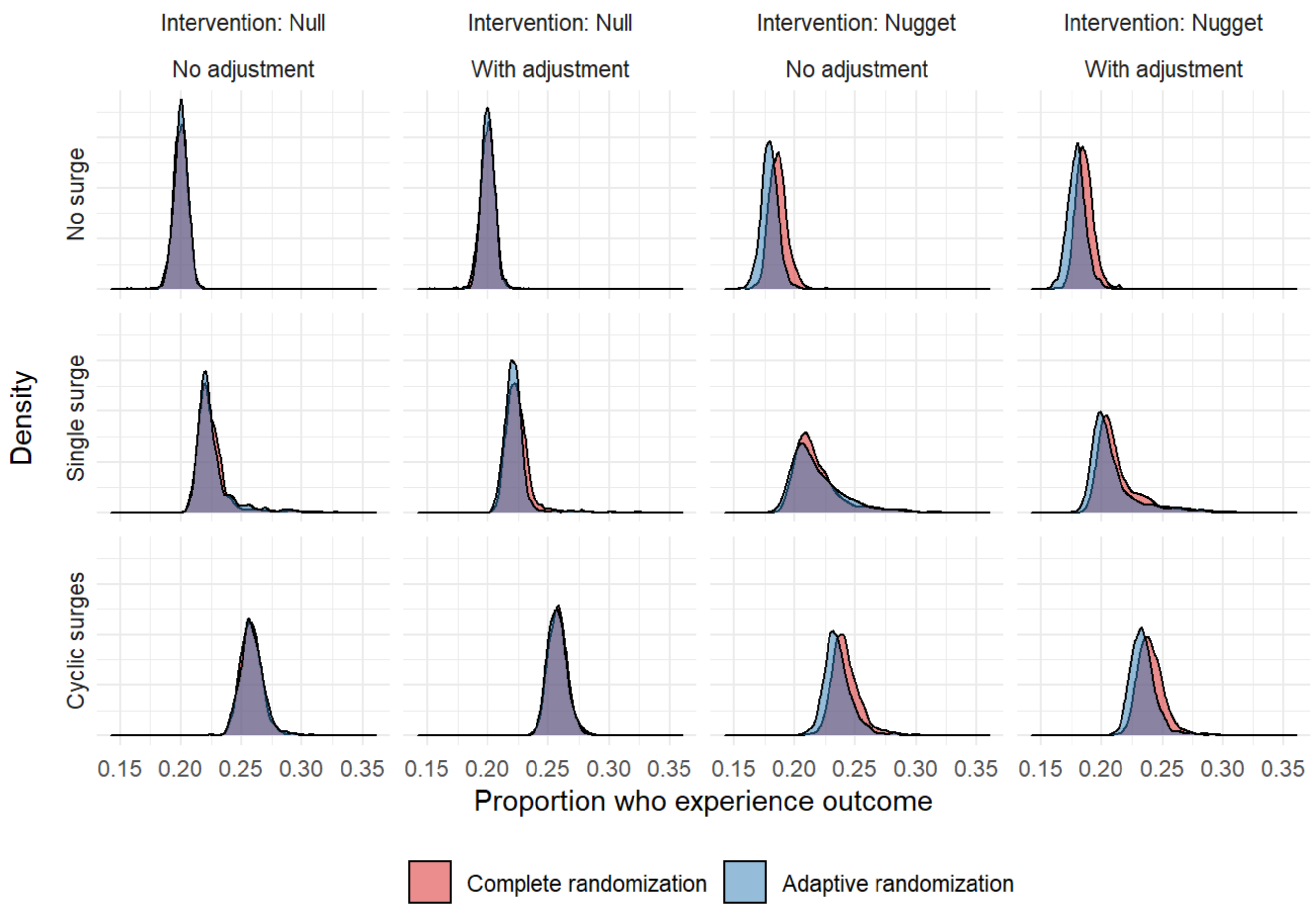

Figure E2 caption: This figure shows the distribution of the proportion patients who died during each trial for each factorial scenario. In null scenarios there is no difference between the randomization algorithms. In nugget scenarios the response-adaptive randomization leads to better outcomes, although the absolute effect is small.

#### Figure E3: Time to dropping intervention 2 or concluding trial according to trial scenario

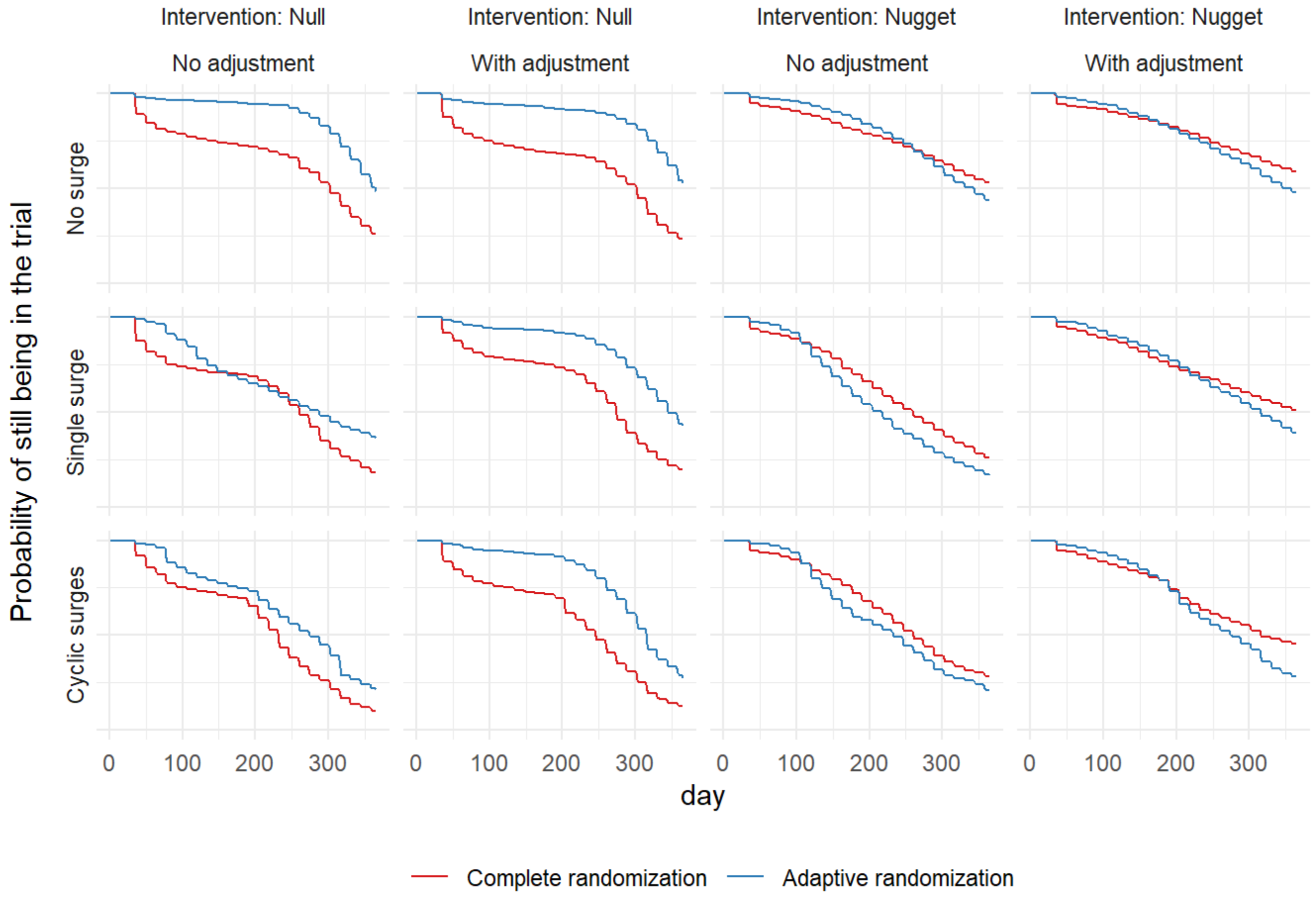

Figure E3 caption: This figure depicts the proportion of trials with intervention 2 remaining active in the trial (y axis) by day of the trial (x axis). In the null scenarios where intervention 2 has OR 1, there is a consistent trend across other simulation variables where trials using complete randomization drop intervention 2 earlier than trials using adaptive randomization. In the nugget scenarios where intervention 2 has OR 0.75, the curves are more similar although there is a consistent pattern where trials using adaptive randomization initially takes longer to identify the superior intervention and then accelerates, surpassing the trials using complete randomization and more frequently identifying intervention 2 as the superior intervention.

#### Results for trials without interim analyses

The principal comparison in this investigation of simulated Bayesian trials is between a conventional multi-arm trial with frequent interim analyses and a response-adaptive multi-arm trial with frequent interim analyses. Very few trials with conventional randomization employ interim analyses with the frequency used in the principal comparison, and so this may dilute the differences between conventional and adaptive randomization. To test this hypothesis, we simulated trials under the extreme assumption of no interim analyses. The trials run for 365 days then analyze their results.

##### Bias

The trials with no interim analyses had less bias than trials using more frequent interim analyses (Table E4). This reinforces that stopping trials for benefit or harm that will introduce at least some bias in the direction of the stopping rule.

##### Operating characteristics

The trials with no interim analyses did not conclude intervention 2 was superior in any of the 3,000 simulations under the null scenario (Table E5). Despite enrolling 25% more patients than trials with interim analyses, the trials reached conclusions less often (Table E6). The highest proportion of trials that reached a conclusion occurred in the nugget scenario with cyclic surges and analyses adjusted for time trends where 63% of trials concluded that an effective intervention was superior. By contrast, adaptive randomization with adjustment for time trends concluded that an effective intervention was superior in 71% of trials.

#### Ethical import

Trials with no interim analyses randomized more than twice as many patients to ineffective arms compared to trials with frequent interim analyses when an effective arm was present (mean 1442 versus 677 across all nugget scenarios). By design no conclusions were reached before 365 days. The average outcomes reflected solely the underlying COVID epidemiology.

#### Conclusion

Independent of the choice to use adaptive versus complete randomization, trials that study therapies for pandemics should employ frequent interim analyses to arrive at conclusions faster, maximize the probability of identifying effective therapies, and minimize wasted resources by dropping ineffective therapies.

### Tables

#### Table E4: Bias for effective and equivalent interventions in trials without interim analyses

| Covid Scenario | Intervention Scenario[a] | Time trends modeled | Algorithm | Mean posterior odds ratio of treatment[b] | Bias (log-odds) | Monte Carlo SE[c] | RMSE[d] |
|---|---|---|---|---|---|---|---|
| No surge | Null | No | Complete | 1 (0.83 to 1.21) | -0.0015 | 0.0030 | 0.096 |
| | | Yes | Complete | 1 (0.83 to 1.21) | -0.0008 | 0.0030 | 0.095 |
| | Nugget | No | Complete | 0.75 (0.62 to 0.92) | -0.0016 | 0.0033 | 0.103 |
| | | Yes | Complete | 0.75 (0.62 to 0.91) | -0.0043 | 0.0032 | 0.100 |
| Single surge | Null | No | Complete | 1 (0.84 to 1.2) | -0.0005 | 0.0028 | 0.090 |
| | | Yes | Complete | 1.01 (0.84 to 1.21) | 0.0020 | 0.0029 | 0.093 |
| | Nugget | No | Complete | 0.76 (0.63 to 0.91) | 0.0034 | 0.0031 | 0.098 |
| | | Yes | Complete | 0.75 (0.62 to 0.91) | -0.0015 | 0.0031 | 0.097 |
| Cyclic surges | Null | No | Complete | 1 (0.85 to 1.18) | -0.0034 | 0.0026 | 0.083 |
| | | Yes | Complete | 1 (0.85 to 1.18) | -0.0013 | 0.0027 | 0.084 |
| | Nugget | No | Complete | 0.76 (0.64 to 0.9) | 0.0086 | 0.0027 | 0.085 |
| | | Yes | Complete | 0.75 (0.63 to 0.89) | -0.0038 | 0.0029 | 0.092 |

#### Table E5: Conclusions for effective and equivalent interventions in trials without interim analyses

| Covid Scenario | Intervention Scenario[a] | Time trends modeled | Algorithm | Trials that reached a conclusion | Type of conclusion | | Type of non-superiority | | | Monte Carlo SE for Rejection |
|---|---|---|---|---|---|---|---|---|---|---|
| | | | | | Rejection | Superiority | Equivalence | Inferiority | Continue | |
| No surge | Null | No | Complete | 0.140 | 1.000 | 0.000 | 0.402 | 0.070 | 0.528 | 0.000 |
| | | Yes | Complete | 0.147 | 1.000 | 0.000 | 0.447 | 0.074 | 0.479 | 0.000 |
| | Nugget | No | Complete | 0.430 | 0.573 | 0.427 | 0.006 | 0.000 | 0.567 | 0.016 |
| | | Yes | Complete | 0.444 | 0.559 | 0.441 | 0.006 | 0.000 | 0.553 | 0.016 |
| Single surge | Null | No | Complete | 0.207 | 1.000 | 0.000 | 0.497 | 0.058 | 0.445 | 0.000 |
| | | Yes | Complete | 0.177 | 1.000 | 0.000 | 0.472 | 0.068 | 0.460 | 0.000 |
| | Nugget | No | Complete | 0.519 | 0.489 | 0.511 | 0.015 | 0.000 | 0.474 | 0.016 |
| | | Yes | Complete | 0.487 | 0.518 | 0.482 | 0.009 | 0.000 | 0.509 | 0.016 |
| Cyclic surges | Null | No | Complete | 0.387 | 1.000 | 0.000 | 0.617 | 0.064 | 0.319 | 0.000 |
| | | Yes | Complete | 0.354 | 1.000 | 0.000 | 0.593 | 0.061 | 0.346 | 0.000 |
| | Nugget | No | Complete | 0.606 | 0.397 | 0.603 | 0.008 | 0.000 | 0.389 | 0.015 |
| | | Yes | Complete | 0.636 | 0.367 | 0.633 | 0.004 | 0.000 | 0.363 | 0.015 |

#### Table E6: Outcomes of trial for effective and equivalent interventions in trials without interim analyses

| Covid Scenario | Intervention Scenario[a] | Time trends modeled | Algorithm | Probability of being optimal | Probability of being equivalent | Sample size: Control arm | Sample size: Intervention 2 | Sample size: Intervention 3 | Sample size: Total | Mortality rate |
|---|---|---|---|---|---|---|---|---|---|---|
| No surge | Null | No | Complete | 0.262 | 0.820 | 1369 | 1369 | 1368 | 5476 | 0.200 |
| | | Yes | Complete | 0.251 | 0.823 | 1369 | 1367 | 1368 | 5471 | 0.200 |
| | Nugget | No | Complete | 0.946 | 0.230 | 1370 | 1369 | 1370 | 5476 | 0.190 |
| | | Yes | Complete | 0.949 | 0.221 | 1369 | 1369 | 1367 | 5474 | 0.189 |
| Single surge | Null | No | Complete | 0.240 | 0.847 | 1423 | 1424 | 1424 | 5695 | 0.219 |
| | | Yes | Complete | 0.248 | 0.836 | 1424 | 1424 | 1423 | 5696 | 0.219 |
| | Nugget | No | Complete | 0.955 | 0.227 | 1422 | 1423 | 1422 | 5690 | 0.208 |
| | | Yes | Complete | 0.960 | 0.216 | 1423 | 1425 | 1425 | 5695 | 0.208 |
| Cyclic surges | Null | No | Complete | 0.264 | 0.880 | 1534 | 1535 | 1534 | 6136 | 0.254 |
| | | Yes | Complete | 0.256 | 0.872 | 1533 | 1531 | 1535 | 6133 | 0.254 |
| | Nugget | No | Complete | 0.973 | 0.212 | 1532 | 1532 | 1533 | 6128 | 0.242 |
| | | Yes | Complete | 0.975 | 0.198 | 1533 | 1535 | 1537 | 6139 | 0.242 |

### Figures

#### Figure E4: Probability that effective and equivalent interventions are optimal

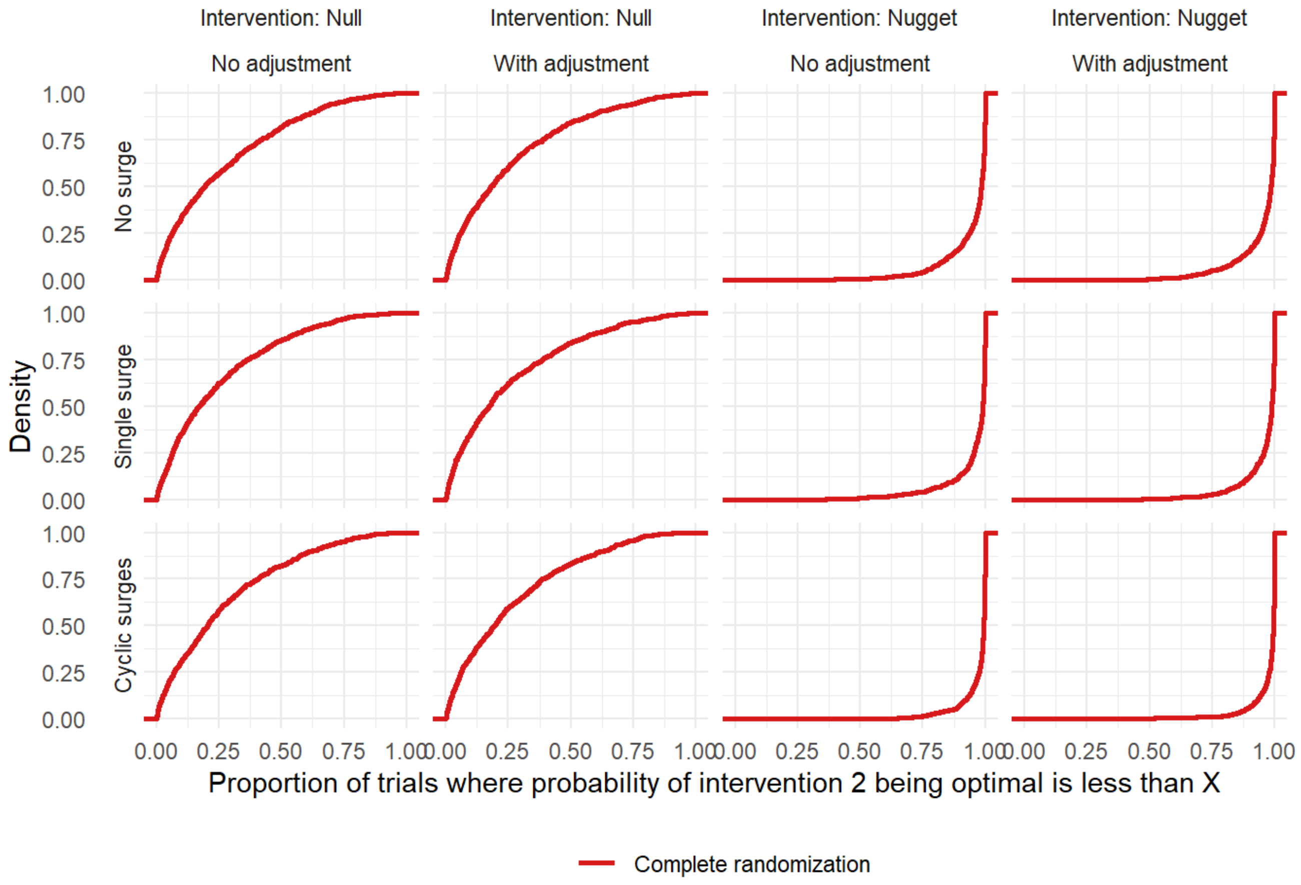

Figure E4 caption: This figure shows the cumulative density distribution of the probabilities that intervention 2 is optimal. For superiority to be declared, the probability that an intervention is optimal must be 0.99 or greater. The figure shows that almost all trials in the null scenario have the probability of intervention 2 being optimal as less than 0.99, and almost all trials in the nugget scenario have the probability of intervention 2 being optimal as greater than 0.95.

#### Results for trials with smaller enrollment rates and time blocks

One pragmatic question for trialists and statisticians is the minimal number of patients in each time block before adjustment for time block can no longer address the bias introduced by pandemic surges. To investigate this question, we considered two additional scenarios with time blocks of 7 instead of 14 days and mean daily enrollment rates of 15 or 6patients per day. This means that there were approximately 50% and 20% as many patients in eachtime block (15*14 = 210 vs 15*7 = 105 vs 6*7 = 42, or expected number of events per time block of 49, 25, or 10). We used the COVID scenario with multiple surges and adjustment for time trends in all trials for this part of the investigation.

##### Bias

The bias results for simulations with a 7-day time block and mean enrollment 15 per day are very similar to the bias results for the analogous trials with 14-day time blocks. However, with smaller enrollment the bias is increased (Table E7). The 95% credible intervals for the means odds ratio in individual trials was wider with lower enrollment, as expected given that the trial duration was still limited to 365 days.

##### Operating characteristics

Reducing the bins until each held only 10 events did not appreciably increase the rate at which equivalent interventions where declared superior (Table E8). The proportion of trials that reached a conclusion in the case with mean enrollment 15 patients per day and time blocks of size 7 was similar to efficiency in analogous trials with time blocks of size 14 (Table E8). This proportion was much diminished with a lower enrollment rate, to the point where if trialists expected the enrollment rate to be only 6 patients per day on average, it would be unreasonable to expect the trial to reach a conclusion in 1 year. However, even among trials with lower enrollment rates that did not finish, if an effective intervention was present the mean across trials of the probability that it was the optimal intervention was greater than 0.80.

#### Ethical import

The two main ethical advantages of adaptive randomization are preserved for scenarios where effective interventions, even at low enrollment rates: the average outcomes remain slightly better and more patients are randomized to the effective intervention. However, at the low enrollment rate, the potential of reaching a conclusion is so low that a trial limited to one year's duration and using the strict stopping rules employed in this investigation would not likely be ethical.

#### Conclusion

For superiority trials targeting mortality in a pandemic, the limiting factor when time blocks have few events is more likely to be low enrollment than bias due to lack of positivity (lack of enough information in each time block). Time blocks of less than one week are impractical, whereas enrollment rates of 6 per day are very plausible if not enough sites are involved in a study. Adaptive randomization, even with adjustment for time trends, cannot rescue a trial that does not enroll enough patients.

### Tables

#### Table E7: Bias for effective and equivalent interventions in trials with 7-day as opposed to 14-day time blocks

| Enrollment rate | Intervention Scenario[a] | Time trends modeled | Algorithm | Mean posterior odds ratio of treatment[b] | Bias (log-odds) | Monte Carlo SE[c] | RMSE[d] |
|---|---|---|---|---|---|---|---|
| 15 daily | Null | Yes | Complete | 1.06 (0.84 to 1.34) | 0.0460 | 0.0045 | 0.151 |
| | | Yes | Adaptive | 1.02 (0.83 to 1.25) | 0.0108 | 0.0040 | 0.127 |
| | Nugget | Yes | Complete | 0.74 (0.61 to 0.91) | -0.0245 | 0.0046 | 0.147 |
| | | Yes | Adaptive | 0.73 (0.59 to 0.91) | -0.0361 | 0.0042 | 0.139 |
| 6 daily | Null | Yes | Complete | 1.08 (0.78 to 1.52) | 0.0530 | 0.0067 | 0.219 |
| | | Yes | Adaptive | 1.05 (0.77 to 1.45) | 0.0270 | 0.0068 | 0.216 |
| | Nugget | Yes | Complete | 0.75 (0.57 to 1.01) | -0.0106 | 0.0057 | 0.182 |
| | | Yes | Adaptive | 0.72 (0.54 to 0.99) | -0.0556 | 0.0063 | 0.208 |

#### Table E8: Conclusions for effective and equivalent interventions in trials with 7- as opposed to 14-day time blocks

| Enrollment rate | Intervention Scenario[a] | Time trends modeled | Algorithm | Trials that reached a conclusion | Type of conclusion | | Type of non-superiority | | | Monte Carlo SE for Rejection |
|---|---|---|---|---|---|---|---|---|---|---|
| | | | | | Rejection | Superiority | Equivalence | Inferiority | Continue | |
| 15 daily | Null | Yes | Complete | 0.787 | 0.996 | 0.004 | 0.555 | 0.325 | 0.116 | 0.002 |
| | | Yes | Adaptive | 0.674 | 0.995 | 0.005 | 0.606 | 0.216 | 0.173 | 0.002 |
| | Nugget | Yes | Complete | 0.595 | 0.473 | 0.527 | 0.017 | 0.056 | 0.400 | 0.016 |
| | | Yes | Adaptive | 0.758 | 0.293 | 0.707 | 0.017 | 0.040 | 0.236 | 0.014 |
| 6 daily | Null | Yes | Complete | 0.062 | 0.993 | 0.007 | 0.013 | 0.303 | 0.677 | 0.003 |
| | | Yes | Adaptive | 0.038 | 0.991 | 0.009 | 0.001 | 0.296 | 0.694 | 0.003 |
| | Nugget | Yes | Complete | 0.194 | 0.828 | 0.172 | 0.002 | 0.053 | 0.773 | 0.012 |
| | | Yes | Adaptive | 0.270 | 0.758 | 0.242 | 0.000 | 0.065 | 0.693 | 0.014 |

#### Table E9: Outcomes of trial for effective and equivalent interventions with 7- as opposed to 14-day time blocks

| Enrollment rate | Intervention Scenario[a] | Time trends modeled | Algorithm | Probability of being optimal | Probability of being equivalent | Sample size: Control arm | Sample size: Intervention 2 | Sample size: Intervention 3 | Sample size: Total | Mortality rate |
|---|---|---|---|---|---|---|---|---|---|---|
| 15 daily | Null | Yes | Complete | 0.250 | 0.753 | 1619 | 1126 | 1113 | 4972 | 0.257 |
| | | Yes | Adaptive | 0.257 | 0.795 | 1445 | 1326 | 1328 | 5388 | 0.257 |
| | Nugget | Yes | Complete | 0.924 | 0.162 | 1696 | 1623 | 683 | 4663 | 0.242 |
| | | Yes | Adaptive | 0.962 | 0.178 | 849 | 2307 | 686 | 4528 | 0.235 |
| 6 daily | Null | Yes | Complete | 0.253 | 0.582 | 741 | 565 | 561 | 2400 | 0.254 |
| | | Yes | Adaptive | 0.238 | 0.583 | 612 | 586 | 592 | 2389 | 0.255 |
| | Nugget | Yes | Complete | 0.827 | 0.250 | 757 | 722 | 407 | 2288 | 0.241 |
| | | Yes | Adaptive | 0.853 | 0.241 | 443 | 972 | 404 | 2213 | 0.235 |

### Figures

#### Figure E5: Bias for trials using time blocks of 7 days and a smaller enrollment rate

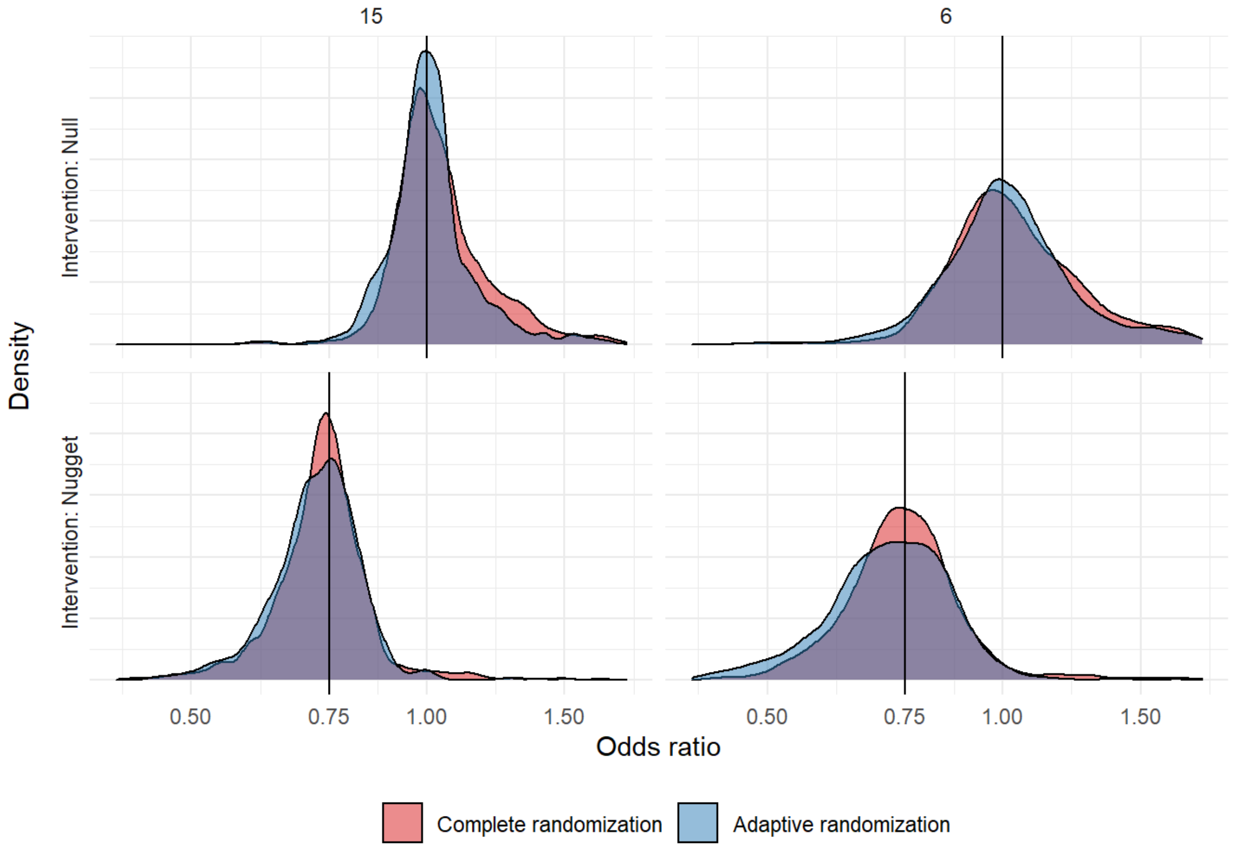

Figure E5 caption: This figure depicts the distribution of odds ratios estimated from simulated trials using cyclic surge epidemiology, adjustments for time trends, time-blocks of size 7 days, either mean daily enrollment rate of 15 (left column) or 6 (right column), null or nugget scenarios (rows), and either complete or adaptive randomization (colours). This figure shows that the lower enrollment rate leads to less information being gathered over the course of the 365-day trial and therefore a more diffuse distribution of odds ratios.

#### Figure E6: Conclusions for trials using time blocks of 7 days and a smaller enrollment rate

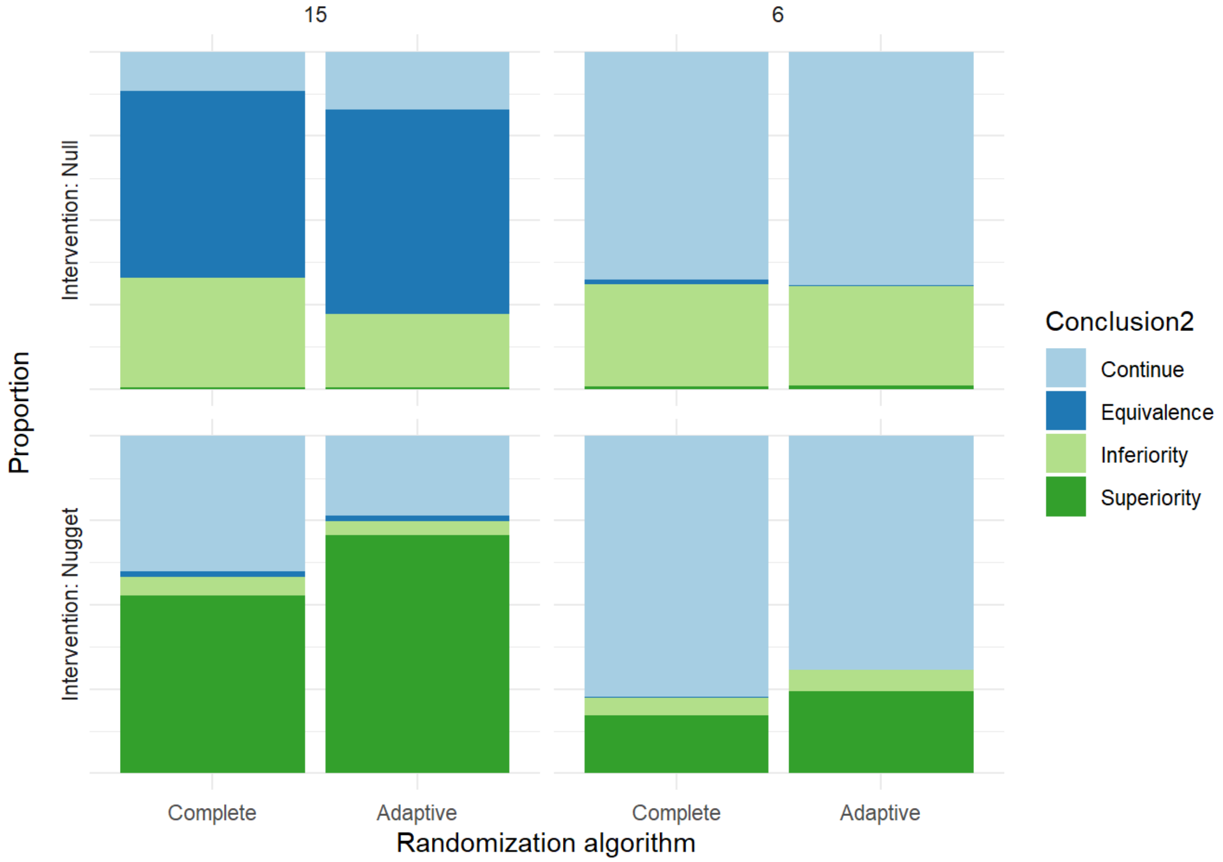

Figure E6 caption: This figure depicts the conclusions for intervention 2 from simulated trials using cyclic surge epidemiology, adjustments for time trends, time-blocks of size 7 days, either mean daily enrollment rate of 15 (left two columns) or 6 (right two columns), null or nugget scenarios (rows), and either complete (columns 1 and 3) or adaptive (columns 2 and 4) randomization. This figure shows that although adaptive randomization continues to identify effective interventions more frequently than complete randomization, the lower enrollment rate dramatically reduces the ability of the trial to reach conclusions.

#### Figure E7: Time to dropping intervention 2 or concluding trial for trials using time blocks of 7 days and a smaller enrollment rate

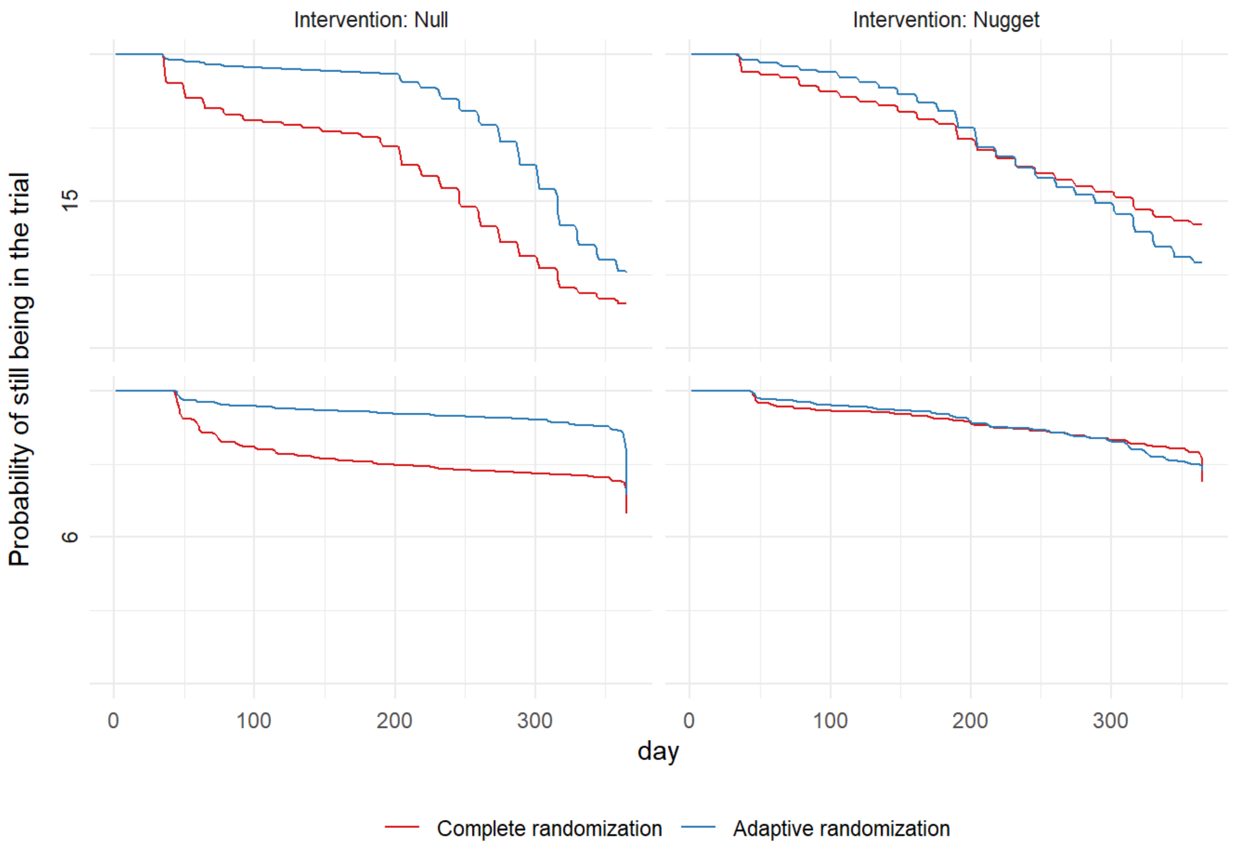

Figure E7 caption: This figure shows the time (in days) to dropping intervention 2 when equivalent (left column, null scenario) or time to conclusion when superior (right column, nugget scenario) according to mean enrollment rate (15, top row; 6, bottom row). The trials have cyclic surge epidemiology, adjust for time trends, and show the randomization method in colour. The lower enrollment rate dramatically reduces the ability of the trial to reach conclusions.

#### Results for trials with later surges

In the base case, the epidemiologic surge occurs relatively early in the trial while allocations are may still be equal because an interim analysis may not occur until midway through the surge. As a sensitivity analysis we also assessed the effects of a later surge on the bias of response-adaptive randomization, using two additional single surge scenarios where the 21-day surge starts on day 150 or on day 270 instead of day 30.

##### Bias

The bias results for mid- and late-trial surges are similar to the results for early-trial surges, with the exception that estimates from trials with mid- and late-trial surges and unadjusted analyses have less bias (Table E10, Figure E7).

##### Operating characteristics

The probability of declaring an equivalent intervention superior remained small, with the largest (1.2%) seen in the late surge scenario with adaptive randomization and adjustment for time trends (Table E11). Trials with mid- and late-trial surges in the nugget scenario had a much lower rate of reaching conclusions when using unadjusted analyses, as opposed to when the surge is early (Table E11). However, the rates of identifying effective interventions were about the same when adjustment for time trends was used. Trials with mid- and late-trial surges using adaptive randomization had a lower rate of identifying effective interventions than trials with early surges, however the advantage that adaptive randomization held over complete randomization in this respect was exaggerated (Figure E8).

#### Ethical import

As in other scenarios, adaptive randomization showed a small mortality benefit across trials with mid- and late-trial surges and complete randomization identified ineffective interventions earlier.

#### Conclusion

When the surge occurs in the middle or near the end of the trial, adjustment for time trends was less important for attenuating bias and more important for increasing the probability of identifying an effective intervention. This provides further evidence that adjustment for time trends should be adopted a priori for trials using adaptive randomization in a pandemic, because it cannot be known at the outset whether a pandemic surge will occur early, late, or not at all.

### Tables

#### Table E10: Bias for effective and equivalent interventions in trials with alternate surge timing

| Covid Scenario | Intervention Scenario[a] | Time trends modeled | Algorithm | Mean posterior odds ratio of treatment[b] | Bias (log-odds) | Monte Carlo SE[c] | RMSE[d] |
|---|---|---|---|---|---|---|---|
| Mid-trial surge | Null | No | Complete | 1.01 (0.8 to 1.28) | 0.0036 | 0.0041 | 0.131 |
| | | | Adaptive | 1.01 (0.83 to 1.24) | 0.0034 | 0.0047 | 0.149 |
| | | Yes | Complete | 1.07 (0.83 to 1.4) | 0.0566 | 0.0054 | 0.178 |
| | | | Adaptive | 1.03 (0.83 to 1.27) | 0.0142 | 0.0045 | 0.142 |
| | Nugget | No | Complete | 0.74 (0.61 to 0.91) | -0.0214 | 0.0045 | 0.145 |
| | | | Adaptive | 0.77 (0.63 to 0.95) | 0.0212 | 0.0044 | 0.142 |
| | | Yes | Complete | 0.74 (0.6 to 0.93) | -0.0210 | 0.0048 | 0.152 |
| | | | Adaptive | 0.72 (0.58 to 0.91) | -0.0464 | 0.0046 | 0.152 |
| Late-trial surge | Null | No | Complete | 1.02 (0.81 to 1.29) | 0.0067 | 0.0041 | 0.130 |
| | | | Adaptive | 1.01 (0.83 to 1.25) | 0.0060 | 0.0039 | 0.125 |
| | | Yes | Complete | 1.08 (0.84 to 1.4) | 0.0623 | 0.0053 | 0.180 |
| | | | Adaptive | 1.03 (0.83 to 1.29) | 0.0167 | 0.0050 | 0.160 |
| | Nugget | No | Complete | 0.75 (0.61 to 0.92) | -0.0136 | 0.0045 | 0.144 |
| | | | Adaptive | 0.77 (0.62 to 0.96) | 0.0155 | 0.0048 | 0.153 |
| | | Yes | Complete | 0.75 (0.61 to 0.93) | -0.0198 | 0.0052 | 0.167 |
| | | | Adaptive | 0.72 (0.58 to 0.91) | -0.0439 | 0.0045 | 0.148 |

#### Table E11: Conclusions for effective and equivalent interventions in trials with alternate surge timing

| Covid Scenario | Intervention Scenario | Time trends modeled | Algorithm | Trials that reached a conclusion | Type of conclusion | | Type of non-superiority | | | Monte Carlo SE for Rejection |
|---|---|---|---|---|---|---|---|---|---|---|
| | | | | | Rejection | Superiority | Equivalence | Inferiority | Continue | |
| Mid-trial surge | Null | No | Complete | 0.744 | 0.996 | 0.004 | 0.510 | 0.368 | 0.118 | 0.002 |
| | | | Adaptive | 0.463 | 0.985 | 0.015 | 0.402 | 0.279 | 0.304 | 0.004 |
| | | Yes | Complete | 0.695 | 0.999 | 0.001 | 0.507 | 0.346 | 0.146 | 0.001 |
| | | | Adaptive | 0.496 | 0.997 | 0.003 | 0.488 | 0.234 | 0.275 | 0.002 |
| | Nugget | No | Complete | 0.420 | 0.651 | 0.349 | 0.013 | 0.061 | 0.577 | 0.015 |
| | | | Adaptive | 0.549 | 0.551 | 0.449 | 0.087 | 0.038 | 0.426 | 0.016 |
| | | Yes | Complete | 0.522 | 0.535 | 0.465 | 0.014 | 0.050 | 0.471 | 0.016 |
| | | | Adaptive | 0.678 | 0.361 | 0.639 | 0.013 | 0.032 | 0.316 | 0.015 |
| Late-trial surge | Null | No | Complete | 0.710 | 0.994 | 0.006 | 0.507 | 0.350 | 0.137 | 0.002 |
| | | | Adaptive | 0.610 | 0.994 | 0.006 | 0.558 | 0.258 | 0.178 | 0.002 |
| | | Yes | Complete | 0.691 | 0.995 | 0.005 | 0.522 | 0.333 | 0.140 | 0.002 |
| | | | Adaptive | 0.488 | 0.988 | 0.012 | 0.499 | 0.236 | 0.253 | 0.003 |
| | Nugget | No | Complete | 0.402 | 0.665 | 0.335 | 0.020 | 0.053 | 0.592 | 0.015 |
| | | | Adaptive | 0.510 | 0.588 | 0.412 | 0.066 | 0.049 | 0.473 | 0.016 |
| | | Yes | Complete | 0.505 | 0.558 | 0.442 | 0.016 | 0.053 | 0.489 | 0.016 |
| | | | Adaptive | 0.668 | 0.390 | 0.610 | 0.021 | 0.050 | 0.319 | 0.015 |

#### Table E12: Outcomes of trial for effective and equivalent interventions in trials with alternate surge timing

| Covid Scenario | Intervention Scenario[1] | Time trends modeled | Algorithm | Probability of being optimal | Probability of being equivalent | Sample size: Control arm | Sample size: Intervention 2 | Sample size: Intervention 3 | Sample size: Total | Mortality rate |
|---|---|---|---|---|---|---|---|---|---|---|
| Mid-trial surge | Null | No | Complete | 0.273 | 0.755 | 1548 | 1053 | 1048 | 4703 | 0.223 |
| | | | Adaptive | 0.258 | 0.725 | 1342 | 1301 | 1269 | 5186 | 0.221 |
| | | Yes | Complete | 0.244 | 0.711 | 1600 | 1092 | 1097 | 4876 | 0.222 |
| | | | Adaptive | 0.254 | 0.771 | 1406 | 1330 | 1290 | 5295 | 0.220 |
| | Nugget | No | Complete | 0.866 | 0.184 | 1771 | 1702 | 688 | 4818 | 0.203 |
| | | | Adaptive | 0.864 | 0.298 | 918 | 2189 | 860 | 4811 | 0.200 |
| | | Yes | Complete | 0.913 | 0.184 | 1629 | 1569 | 667 | 4539 | 0.205 |
| | | | Adaptive | 0.961 | 0.179 | 824 | 2217 | 690 | 4431 | 0.200 |
| Late-trial surge | Null | No | Complete | 0.258 | 0.758 | 1596 | 1083 | 1113 | 4928 | 0.217 |
| | | | Adaptive | 0.243 | 0.790 | 1365 | 1241 | 1292 | 5175 | 0.219 |
| | | Yes | Complete | 0.237 | 0.715 | 1626 | 1132 | 1115 | 5040 | 0.217 |
| | | | Adaptive | 0.260 | 0.764 | 1404 | 1310 | 1312 | 5323 | 0.219 |
| | Nugget | No | Complete | 0.818 | 0.199 | 1696 | 1628 | 703 | 4702 | 0.199 |
| | | | Adaptive | 0.867 | 0.319 | 860 | 2185 | 746 | 4539 | 0.191 |
| | | Yes | Complete | 0.900 | 0.179 | 1674 | 1599 | 719 | 4717 | 0.200 |
| | | | Adaptive | 0.961 | 0.181 | 849 | 2280 | 714 | 4560 | 0.192 |

### Figures

#### Figure E8: Bias for trials with alternate surge timing

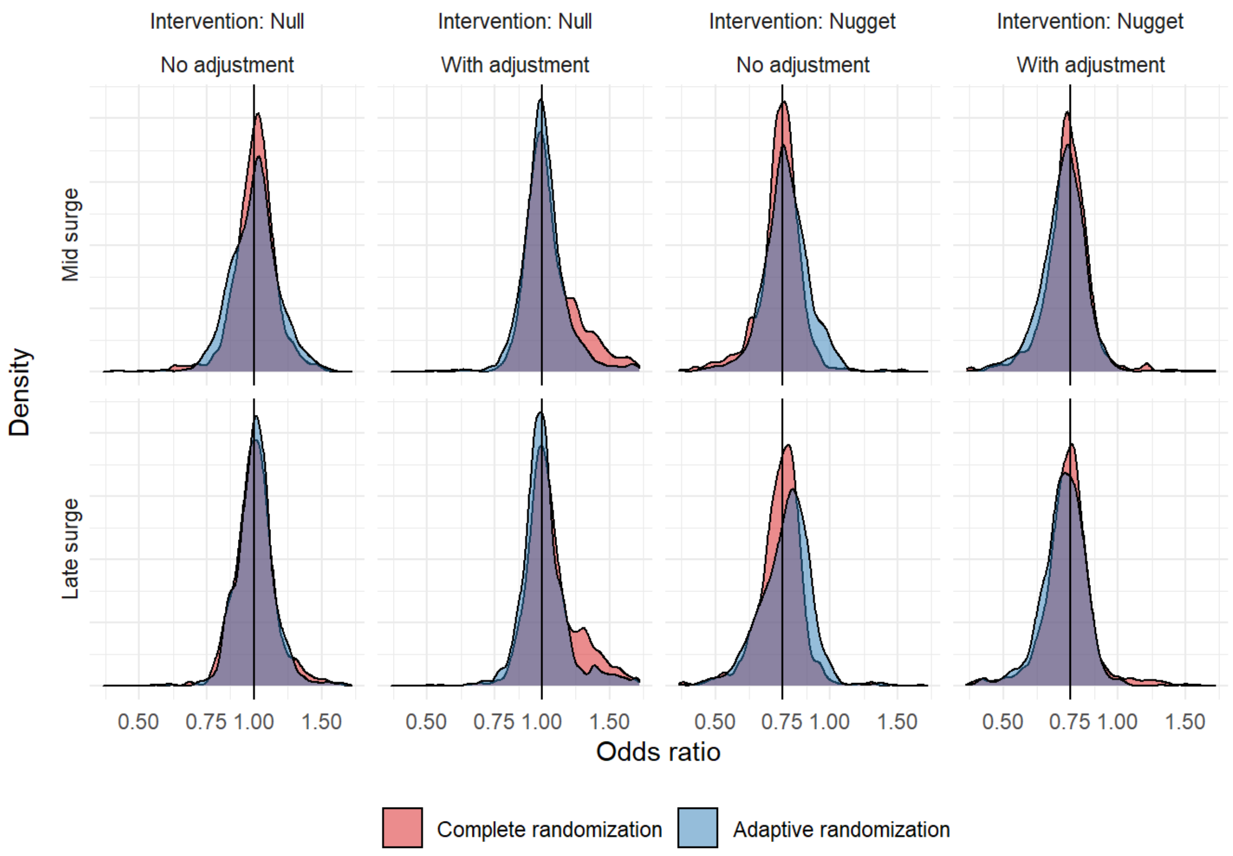

Figure E8 caption: This figure depicts the distribution of odds ratios estimated from simulated trials using alternate surge epidemiology. This figure shows that adaptive randomization without time-trends adjustment leads to less bias when surges occur in the middle or end of the trial, as opposed to when they occur early. As before, adjustment for time trends attenuates the bias observed using adaptive randomization.

#### Figure E9: Conclusions for trials with alternate surge timing

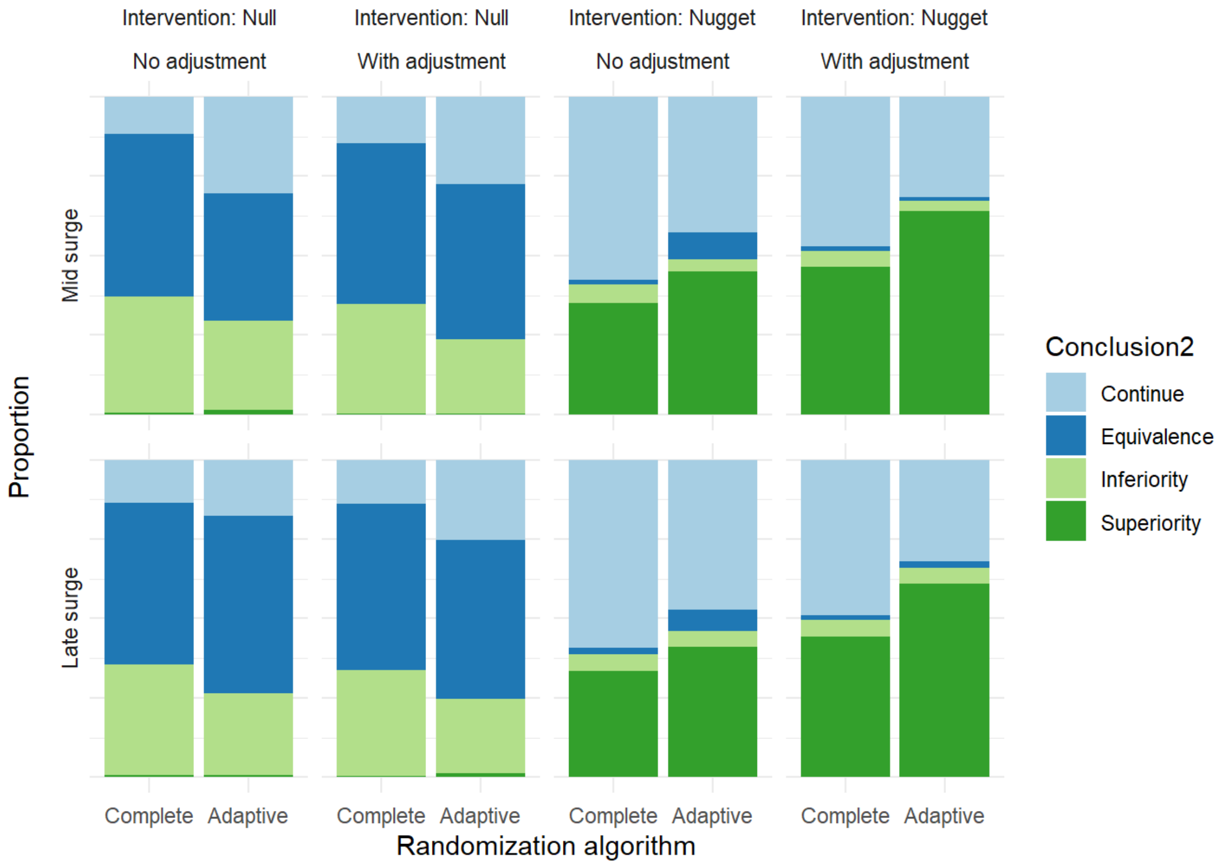

Figure E9 caption: This figure depicts the conclusions for intervention 2 from simulated trials using alternate surge timing. This figure shows that adaptive randomization continues to identify effective interventions more frequently than complete randomization, especially with adjustment for time trends.

#### Figure E10: Time to dropping an equivalent intervention or concluding trial for trials with alternate surge timing

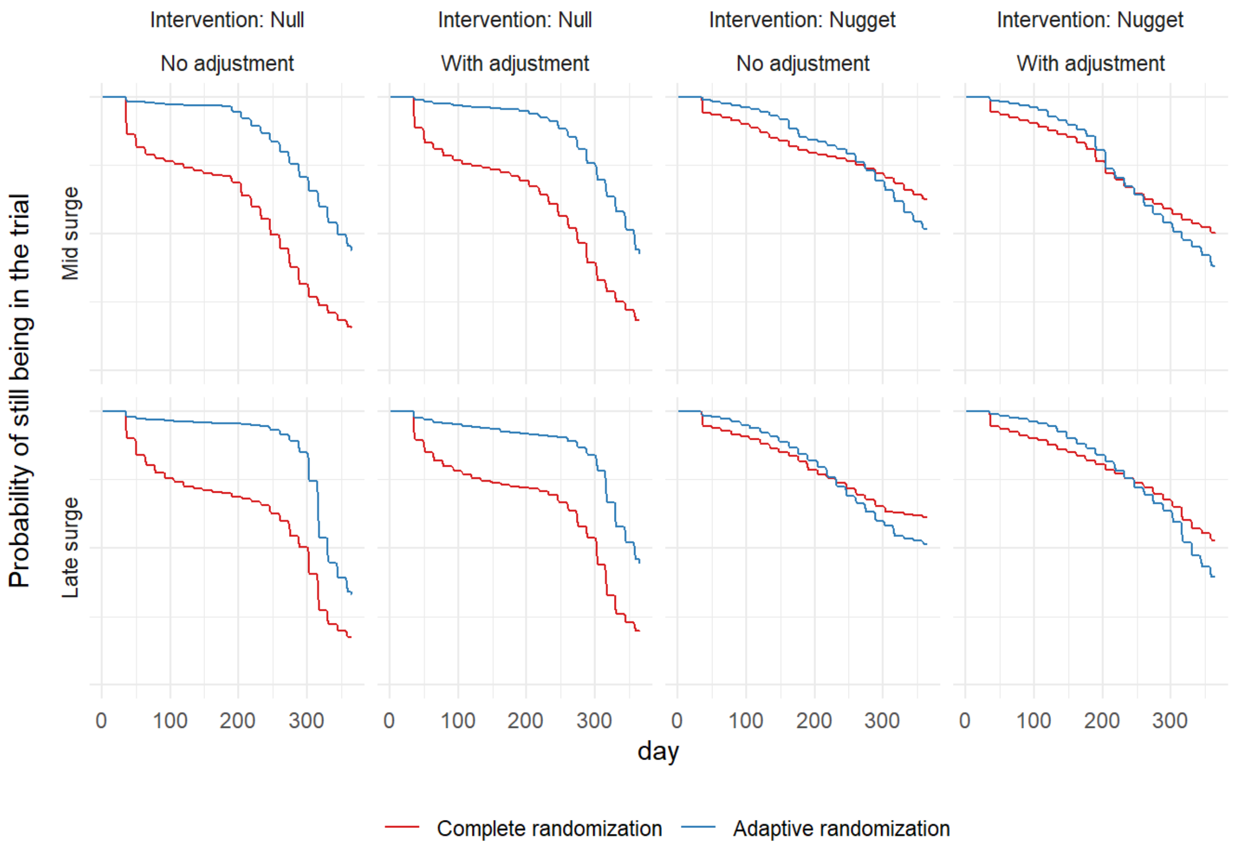

Figure E10 caption: This figure shows the time (in days) to dropping intervention 2 when equivalent or time to conclusion when superior for alternate surge epidemiology. The lower enrollment rate dramatically reduces the ability of the trial to reach conclusions. The rate of conclusions seems to increase during or shortly after the time of a surge. The behavior of the trials, including complete randomization generally dropping ineffective arms earlier, is preserved across different surge locations.

#### Computer code in R

Output below is from RStudio:

```
### PandemicSim Cluster Script

# Table of Contents:

# Package dependencies
# Worker code
  # 0. Parameters
  # 1-9. Functions
  # 10. Base models for stan / brms
# Head node code

##############################################################################
#

# Package dependencies:
# tidyverse
# brms
# parallel

library(dplyr)
library(brms)
library(doParallel)
library(foreach)
library(doRNG)

global.seed <- 1882 # for reproducibility
prefix <- paste0("PandemicSim",global.seed) # prefix for saving file

##############################################################################
#

# Worker code:

# here we define the baseline parameters and functions
# that will run on all the cluster nodes

##############################################################################
#

# 0. Parameters for the data-generating mechanism

cfr <- c(0.2, 0.4) # control group mortality rate for baseline and surge cond
itions

er <- c(15, 25) # mean enrollment rates for baseline and surge conditions
```

```
arms <- 3 # number of arms in addition to control

#odds ratios for interventions
effective <- 0.75

covid_params <- data.frame("cfr" = cfr,
                           "surge" = c(F, T),
                           "er"   = er)

# each column corresponds to a different intervention scenario
intervention_params <- data.frame("NoEffects" = rep(1,arms),
                                  "OneWorks"  = c(effective, rep(1, arms-1)))

allo <- c("CR", "RC_TS") # randomization algorithm, TyRo also an option

nsim <- 1000  # number of simulations per factorial scenario

interim_interval <- 14 # interval between interim analyses in days
burnin_n <- 100 # number of patients to enroll with completed outcome before
first interim analysis
outcome_delay <- 28 # days after enrollment to know the outcome
time_block <- 2*7 # size of time blocks for analysis in days

thresholds <- c(0.99, 0.01, 0.90) # threshold posterior probability for super
iority /inferiority / equivalence

rope <- c(1/1.2, 1.2)

days <- 365 # maximum duration of the trial

surge_date   <- 30 # how many days into the simulation is the surge
surge_length <- 21 # how many days long is the surge
surge_cycle <- 120 # days between surges

covid_scenario_order <- c(1,2,3) # 1, 2, or 3:
# 1 = no surge
# 2 = one surge
# 3 = cyclic surges

intervention_scenario_order <- c("NoEffects","OneWorks")
# recall that "NoEffects", "OneWorks", "OneHarms", and "OneWOneH" are the opt
ions

model_time_trends_order <- c(F,T)

##############################################################################
```

```
#

# 1. expit for inverting odds ratios to probabilities

expit <- function(x){exp(x)/(1+exp(x))}

# 2. enrollment_rate for calculating enrollment rate as a function of covid e
pidemiology and day of trial

enrollment_rate <- function(day, covid_params,
                            covid_scenario, surge_status){
  err <- filter(covid_params, surge == surge_status)$er
}

# 3. CI95 for calculating 95% credibility intervals

CI95 <- function(x){quantile(x, c(0.025,0.975))}

# 4. Interim analysis function

interimRAR <- function(x, r_allo, allocate, outcomes_available,
                       day, thresholds, rope, trial_status,
                       model_time_trends, trial_model, time_block){

  x2 <- filter(x, enrolldate < outcomes_available & enrolldate > 0)

  if (model_time_trends){x2 <- mark_time_blocks(x2, time_block)}

  # bayesian analysis, outputs a list with vector of posterior odds ratios re
lative to control and posterior probabilities of the outcome
  results <- trial_analysis(data = x2,
                            model = trial_model,
                            model_time_trends = model_time_trends)

  post_equiv <- apply(((results[["post_or"]] > rope[1]) & (results[["post_or"
]] < rope[2])), 2, mean)

  p_k <- apply(results[["post_prob"]],2,mean)

  # which intervention has the lowest outcome rate (which intervention is bes
t) for each MCMC iteration
  mins <- factor(apply(results[["post_prob"]], 1, which.min), levels = 1:(arm
s+1))

  # convert to probabilities
  opti  <- table(mins)/length(mins)

  # interim analysis for REMAP-CAP style Thompson sampling, a myopic patient-
benefit focused algorithm
  if(r_allo == "RC_TS"){
```

```
    m_k <- count(x2, treatment)$n # number of patients in each group so far
    rho <- sqrt(opti/(1+m_k))
    allocate <- rho/sum(rho)

    # similar to REMAP-CAP, if new allocation probability is less than 0.1 we
set it to 0.1 and renormalize
    # in fact they do something slightly more sophisticated - this will resul
t in new allocation probabilities slightly
    # less than 0.1 and in REMAP CAP they add a nuisance parameter to the odd
s ratio so that it works out
    # it doesn't make a big difference here
    which_leq_0.1 <- which(allocate < 0.1)
    nuisance <- 0.1-allocate[which_leq_0.1]
    allocate[-which_leq_0.1] <- allocate[-which_leq_0.1]*(1-sum(nuisance))
    allocate[which_leq_0.1] <- 0.1
    allocate <- allocate/sum(allocate) # just in case rounding caused problem
s
  }

  # Tymofyeyev and Rosenberger method is myopic power-oriented
  if(r_allo == "TyRo"){

    # convert to log-odds of mortality for each of the arms
    # because we need the probability of outcome in each arm
    p_k2 <- 1-p_k # median posterior probability of successful outcome
    N <- length(x2$treatment) # number of patients enrolled so far (with outc
omes available)
    n_k <- count(x2, treatment)$n/N # vector for the proportion of patients e
nrolled in each arm so far

    gamma <- 2 # constant that can be titrated to adjust the algorithm; at 0
it is complete randomization and at infinity it is deterministic

    rho <- p_k2*(p_k2/n_k)^gamma

    allocate <- rho/sum(rho)

  }

  # interim analysis using classical randomization / group sequential design
needs no extra algorithm

  # check stopping rules and adjust allocation accordingly

  trial_status[opti[-1]>thresholds[1]] <- "Superiority"

  trial_status[post_equiv > thresholds[3]] <- "Equivalence"
  allocate[-1][post_equiv > thresholds[3]] <- 0
```

```
  trial_status[opti[-1]<thresholds[2]] <- "Inferiority"
  allocate[-1][opti[-1]<thresholds[2]] <- 0 # drop inferior arm

  allocate <- allocate/sum(allocate)

  list("allocate" = allocate,
       "trial_status" = trial_status,
       "ProbOpt2" = opti[2],
       "ProbOpt3" = opti[3],
       "ProbOpt4" = opti[4],
       "ProbEquiv1" = post_equiv[1],
       "ProbEquiv2" = post_equiv[2],
       "ProbEquiv3" = post_equiv[3],
       "Allo1" = allocate[1],
       "Allo2" = allocate[2],
       "Allo3" = allocate[3],
       "Allo4" = allocate[4])
}

# 5. mark_time_blocks for adding the time block variable to the trial data

mark_time_blocks <- function(x, time_block){
  max_day <- max(x[,1])
  extra_days <- max_day %% time_block
  blocks <- (max_day-extra_days)/time_block
  breaks <-  c(0, seq(1:blocks)*time_block + extra_days)
  mutate(x,
         time_block = cut(enrolldate,
                          breaks = breaks,
                          labels = 1:blocks))
}

# 6. final for performing the final analysis on the dataset after trial concl
usion

final <- function(x, time_block, day, trial_status, model_time_trends, trial_
model){

  if (model_time_trends){x <- mark_time_blocks(x, time_block)}

  results <- trial_analysis(data = x,
                            model = trial_model,
                            model_time_trends = model_time_trends)

  post_equiv <- apply(((results[["post_or"]] > rope[1]) & (results[["post_or"
]] < rope[2])), 2, mean)

  p_k <- apply(results[["post_prob"]],2,mean)
```

```
  # which intervention has the lowest outcome rate (which intervention is bes
t) for each MCMC iteration
  mins <- factor(apply(results[["post_prob"]], 1, which.min), levels = 1:(arm
s+1))

  # convert to probabilities
  opti  <- table(mins)/length(mins)

  # observed outcome
  observed_outcome <- mean(x$outcome)

  # trial conclusions:  # opti[-1] drops the first item from opti vector (con
trol)
  trial_status[opti[-1]>thresholds[1]] <- "Superiority"
  trial_status[post_equiv > thresholds[3]] <- "Equivalence"
  trial_status[opti[-1]<thresholds[2]] <- "Inferiority"

  list(exp(mean(results[["post_logodds"]][,1])),
       exp(quantile(results[["post_logodds"]][,1],0.025)),
       exp(quantile(results[["post_logodds"]][,1],0.975)),
       exp(mean(results[["post_logodds"]][,2])),
       exp(quantile(results[["post_logodds"]][,2],0.025)),
       exp(quantile(results[["post_logodds"]][,2],0.975)),
       exp(mean(results[["post_logodds"]][,3])),
       exp(quantile(results[["post_logodds"]][,3],0.025)),
       exp(quantile(results[["post_logodds"]][,3],0.975)),
       opti[1],
       opti[2],
       opti[3],
       opti[4],
       post_equiv[1],
       post_equiv[2],
       post_equiv[3],
       trial_status[1],
       trial_status[2],
       trial_status[3],
       observed_outcome
  )
}

# 7. trial_analysis function for performing the Bayesian analysis

trial_analysis <- function(data, model, model_time_trends){

  if(model_time_trends & length(unique(data$time_block)) > 1)
    {test <- update(model[[2]],  newdata = data,
                    chains = 2,
                    recompile = F,
                    silent = T,
                    refresh = 0,
```

```
                    iter = 3000,
                    warmup = 1000,
                    open_progress = F)}
  else
    {test <- update(model[[1]],  newdata = data,
                   chains = 2,
                   recompile = F,
                   silent = T,
                   refresh = 0,
                   iter = 3000,
                   warmup = 1000,
                   open_progress = F)}

  test2 <- posterior_samples(test, pars = c("b_Intercept",
                                            "b_treatment2",
                                            "b_treatment3",
                                            "b_treatment4"))

  post_logodds <- data.frame(trt1 = test2$b_treatment2,
                             trt2 = test2$b_treatment3,
                             trt3 = test2$b_treatment4)

  post_or <- exp(post_logodds)

  post_prob <- data.frame(m1 = expit(test2$b_Intercept),
                          m2 = expit(test2$b_Intercept + test2$b_treatment2),
                          m3 = expit(test2$b_Intercept + test2$b_treatment3),
                          m4 = expit(test2$b_Intercept + test2$b_treatment4))

  list(post_or = post_or,
       post_prob = post_prob,
       post_logodds = post_logodds)

}

# 8. are_we_surged function for identifying surge status as a function of cov
id epidemiology and date of trial

are_we_surged <- function(day, surge_date, surge_length,
                          surge_cycle, covid_scenario){
  if (covid_scenario == 2 &
      day %in% (surge_date:(surge_date+surge_length))){
    T
  } else if (covid_scenario == 3 &
             (day %% surge_cycle) %in% (surge_date:(surge_date+surge_length))
){
    T
  } else {F}
}
```

```r
# 9. outcome_probabilities

# function that calculates the outcome probabilities by treatment group given
covid epidemiology status, intervention parameteres, and day of the trial
outcome_probabilities <- function(day, covid_params,
                                  covid_scenario, intervention_params,
                                  intervention_scenario,
                                  surge_status){

    tempcfr <- filter(covid_params, surge == surge_status)$cfr
    outodds <- c(tempcfr/(1-tempcfr),
                 (tempcfr/(1-tempcfr))*(intervention_params[[intervention_sce
nario]]))
    outodds/(1+outodds)
}

# 10. simulate_trial function which will be fed to the cluster and is the uni
t of parallelization

# this function takes a list a with elements, in order:
# arms, r_allo, outcome_delay,
# burnin_n, covid_params, covid_scenario,
# intervention_params, intervention_scenario,
# time_block, thresholds, rope, days,
# model_time_trends, trial_model

simulate_trial <- function(a){

  arms <- a[[1]]
  r_allo <- a[[2]]
  outcome_delay <- a[[3]]
  burnin_n <- a[[4]]
  covid_params <- data.frame(cfr = a[[5]],
                             surge = a[[6]],
                             er = a[[7]])
  covid_scenario <- a[[8]]
  intervention_params <- data.frame(NoEffects = a[[9]],
                                    OneWorks = a[[10]])
  intervention_scenario <- a[[11]]
  time_block <- a[[12]]
  thresholds <- c(a[[13]], a[[14]], a[[15]])
  rope <- c(a[[16]], a[[17]])
  days <- a[[18]]
  model_time_trends <- a[[19]]
  trial_model <- a[20:21]

  nmax <- 10000 # number of potential patients, makes a row for each in the d
ataframe
```

```
  trial_status = c("Continue","Continue","Continue")

  # initialize dataframe for data collection
  trial_data <- data.frame(
    enrolldate = rep(0,nmax),
    treatment = rep(1,nmax),
    outcome = rep(0,nmax)
  ) %>%
    mutate(treatment = factor(treatment, levels = c(1,2,3,4)))

  allocate <- rep(1/(arms+1), arms+1) # start with complete randomization
  new_pts  <- 0
  data_counter <- 1
  first_interim <- 0

  for (i in 1:days){

    # Do interim analysis if at least X patients and on an interim analysis t
ime
    if(
      # do first interim analysis after observing outcomes on burnin_n patien
ts
      (first_interim == 0) &
      (sum((trial_data$enrolldate > 0) & (trial_data$enrolldate < i-outcome_d
elay)) > burnin_n)
    ){
      first_interim <- i
      interim_analysis <- interimRAR(
        filter(trial_data, enrolldate > 0),
        r_allo = r_allo,
        allocate = allocate,
        outcomes_available = i-outcome_delay,
        day = i,
        thresholds = thresholds,
        rope = rope,
        trial_status = trial_status,
        model_time_trends = model_time_trends,
        trial_model = trial_model,
        time_block = time_block)

      allocate <- interim_analysis[[1]][]

      trial_status <- interim_analysis[[2]][]
    }
    if(
      # do interim analysis at interim_interval intervals after the first int
erim analysis
      (first_interim > 0) &
      ((i-first_interim) %% interim_interval == 0)){
      interim_analysis <- interimRAR(
```

```
        filter(trial_data, enrolldate > 0),
        r_allo = r_allo,
        allocate = allocate,
        outcomes_available = i-outcome_delay,
        day = i,
        thresholds = thresholds,
        rope = rope,
        trial_status = trial_status,
        model_time_trends = model_time_trends,
        trial_model = trial_model,
        time_block = time_block)

      allocate <- interim_analysis[[1]][]

      trial_status <- interim_analysis[[2]][]
    }

    # check if stopping rules met

    if("Superiority" %in% trial_status){break} # stop if superiority proven
    if(!("Continue" %in% trial_status)){break} # stop if no arms are in "Cont
inue"

    # check surge status
    surge_status <- are_we_surged(day = i,
                                  surge_date,
                                  surge_length,
                                  surge_cycle,
                                  covid_scenario)

    # find enrollment rate as function of surge status
    err <- enrollment_rate(day = i, covid_params = covid_params,
                           covid_scenario = covid_scenario,
                           surge_status = surge_status)

    # enroll new patients
    new_pts <- rpois(1,err)

    if (new_pts > 0){
      # randomize according to current allocation probabilities
      treatments <- sample(arms+1, new_pts, prob = allocate, replace = T)

      # find outcomes as a function of allocation and surge status
      probs <- outcome_probabilities(day = i, covid_params = covid_params,
                                     covid_scenario = covid_scenario,
                                     intervention_params = intervention_param
s,
                                     intervention_scenario = intervention_sce
nario,
                                     surge_status = surge_status)
```

```r
    # generate outcomes according to treatment allocation
    outcomes <- rbinom(n = new_pts, size = 1, prob = probs[treatments])

    trial_data[data_counter:(data_counter+new_pts-1),] <-
      matrix(c(rep(i,new_pts), treatments, outcomes),ncol=3)

    data_counter <- data_counter + new_pts
  }
}

final_data <- filter(trial_data, enrolldate > 0)

# analyze all outcome data (no delay)
final_analysis <- final(final_data,
                        time_block = time_block,
                        day = i,
                        trial_status = trial_status,
                        model_time_trends = model_time_trends,
                        trial_model = trial_model)

# hardwired for 3 arms in addition to control at present
    c(as.numeric(final_analysis[[1]]),
  as.numeric(final_analysis[[2]]),
  as.numeric(final_analysis[[3]]),
  as.numeric(final_analysis[[4]]),
  as.numeric(final_analysis[[5]]),
  as.numeric(final_analysis[[6]]),
  as.numeric(final_analysis[[7]]),
  as.numeric(final_analysis[[8]]),
  as.numeric(final_analysis[[9]]),
  as.numeric(final_analysis[[10]]),
  as.numeric(final_analysis[[11]]),
  as.numeric(final_analysis[[12]]),
  as.numeric(final_analysis[[13]]),
  as.numeric(final_analysis[[14]]),
  as.numeric(final_analysis[[15]]),
  as.numeric(final_analysis[[16]]),
  final_analysis[[17]],
  final_analysis[[18]],
  final_analysis[[19]],
  as.numeric(sum(final_data$treatment == 1)),
  as.numeric(sum(final_data$treatment == 2)),
  as.numeric(sum(final_data$treatment == 3)),
  as.numeric(sum(final_data$treatment == 4)),
  as.numeric(max(final_data$enrolldate)),
  as.numeric(max(filter(final_data, treatment == 1)$enrolldate)),
  as.numeric(max(filter(final_data, treatment == 2)$enrolldate)),
  as.numeric(max(filter(final_data, treatment == 3)$enrolldate)),
  as.numeric(max(filter(final_data, treatment == 4)$enrolldate)),
```

```
    as.numeric(final_analysis[[20]]),
    covid_scenario,
    intervention_scenario,
    r_allo,
    model_time_trends)
}

# 10. Base models for stan / brms

# base models for stan / brms

trial_model <- readRDS("trial_model.rds")

trial_model_time <- readRDS("trial_model_time.rds")

trial_models <- list(trial_model,
                     trial_model_time)

##############################################################################
#
# parallelization code
#

# generate list of parameter combinations, one entry for each simulated trial

indexlist <- list()
index <- 1

for (m in 1:length(covid_scenario_order)){

  covid_scenario <- covid_scenario_order[m]

  for (n in 1:length(intervention_scenario_order)){

    intervention_scenario <- intervention_scenario_order[n]

    for (k in 1:length(allo)){

      r_allo <- allo[k]

      for (i in 1:length(model_time_trends_order)){

        model_time_trends <- model_time_trends_order[i]

        for (j in 1:nsim){

          indexlist[index] <- list(
            c(arms, r_allo, outcome_delay,
              burnin_n, covid_params,
```

```r
              covid_scenario,
              intervention_params,
              intervention_scenario,
              time_block,
              thresholds, rope,
              days,
              model_time_trends,
              trial_models)
          )
          index <- index + 1

        }
      }
    }
  }
}

# Use the environment variable SLURM_CPUS_PER_TASK to set the number of cores
.
# This is for SLURM. Replace SLURM_CPUS_PER_TASK by the proper variable for y
our system.
# Avoid manually setting a number of cores.
ncores = Sys.getenv("SLURM_CPUS_PER_TASK")

registerDoParallel(cores=ncores)# Shows the number of Parallel Workers to be
used
print(ncores) # this how many cores are available, and how many you have requ
ested.
getDoParWorkers()# you can compare with the number of actual workers
registerDoRNG(global.seed) # sets the random number seed appropriately for pa
rallelization

result <- foreach(a = 1:length(indexlist)) %dopar% {simulate_trial(indexlist[
[a]])}

filename <- paste(prefix, "_", Sys.Date(), ".rds", sep = "")
saveRDS(result, file = filename)
```

21. Viele K, Broglio K, McGlothlin A, Saville BR. Comparison of methods for control allocation in multiple arm studies using response adaptive randomization. *Clin TRIALS*.

2020;17(1):52-60. doi:10.1177/1740774519877836
